# Prevalence of Clinically Actionable Cerebrospinal Fluid Abnormalities in First Episode Psychosis: A Systematic Review and Meta-analysis

**DOI:** 10.1101/2025.10.22.25338558

**Authors:** Anjali Chandra, Laura Duque, Andrew Pines, Anne Fladger, Giovanna S. Manzano, Michael E. Benros, Graham Blackman, Matthew L. Baum

## Abstract

**Importance:** First-episode psychosis (FEP) may result from a variety of secondary etiologies, making lumbar puncture (LP) for cerebrospinal fluid (CSF) investigations an important diagnostic consideration in this context. However, the lack of high-quality evidence on the prevalence of clinically-relevant CSF abnormalities hampers clinical consensus on when to pursue LP in FEP.

**Objective:** Determine a meta-analytic estimate of the prevalence of clinically-relevant CSF abnormalities in FEP.

**Data Sources:** Electronic databases Ovid, Medline, Embase, PsychoINFO, and Web of Science were searched from inception to October, 2024. References of included articles were also screened.

**Study Selection:** We included studies that performed LP on a cohort of FEP and reported results of clinically available CSF analysis, enabling prevalence-estimates of abnormalities.

**Data Extraction and Synthesis:** Data was extracted following PRISMA and MOOSE guidelines. Pooled prevalences were calculated by random-effects models. Moderators were tested using meta-regression analysis, and heterogeneity assessed by I^2^ index.

**Main Outcomes and Measures:** Prevalence of CSF abnormalities, focusing on clinically-relevant markers and number needed to assess (NNA).

**Results:** Thirty-seven papers comprising 3,330 FEP patients who underwent LP were included, allowing calculation of prevalence-estimates for 12 CSF abnormalities. Of clinically-relevant abnormalities, the prevalence of CSF-restricted oligoclonal bands (OCB2) was 7.1% (95% CI 3.3-12.0, NNA 14), pleocytosis was 3.2% (95% CI 2.1-4.4, NNA 31) and anti-NMDAR antibodies was 2.3% (95% CI 0.1-6.4, NNA 43). Subgroup analysis showed that anti-neuronal antibodies were mainly detected in studies that selected patients with high suspicion of secondary causes and were low in studies which excluded patients with a high index of suspicion of a secondary cause, based on clinical and ancillary testing. OCBs and pleocytosis also had higher prevalence in the high-suspicion subgroup but were still detected at prevalence even in the low-suspicion subgroup.

**Conclusions and Relevance:** The meta-analytic estimate of the prevalence of the most common clinically relevant CSF abnormality was 7.1%, which is similar to the prevalence of finding any clinically-relevant radiologic abnormality with an MRI brain. Subgroup-analysis supports the usefulness of methods to estimate the pre-LP probability of clinically-relevant CSF abnormalities, albeit these methods are better applied for some abnormalities (CNS-reactive antibodies) than others (OCB2).

**Key Points:** *Question:* What is the prevalence of clinically actionable cerebrospinal fluid abnormalities in first-episode psychosis (FEP)?

*Findings:* In this systematic review and meta-analysis including 3330 patients with FEP who systematically received a lumbar puncture, at least 7.1% had a clinically-relevant abnormality detected in the cerebrospinal fluid; prevalence of oligoclonal bands was 7.1%, pleocytosis was 3.2%, anti-NMDAR antibodies were 2.3%, and other anti-neuronal antibodies were 0.4%. Subgroup-analysis identified association between prior suspicion of secondary causes and prevalence-estimates.

*Meaning:* Lumbar puncture detects clinically-relevant abnormalities in FEP at a similar rate to brain MRI and may be especially informative when there is heightened suspicion for secondary psychosis.

## Introduction

Approximately 3 to 4 out of 100 people will experience a first episode of psychosis (FEP) in their lifetime^1^. Careful clinical assessment and use of ancillary diagnostic procedures is used to identify the subgroup of these cases that have secondary causes, including toxic, neurologic, metabolic, or autoimmune dysfunction, which carry distinct treatment and prognostic implications.^2^ Because some causes of secondary psychosis may yield detectable abnormalities in the cerebrospinal fluid, lumbar puncture (LP) is sometimes considered in the evaluation of FEP. Lumbar puncture (LP) is a well-established general medical procedure with a high sensitivity and specificity for detection of neuroinflammation ^3–6^, but its clinical utility remains uncertain for the evaluation of FEP. While many posit that LP should be restricted to cases where there exists a high suspicion of neuroinflammation (i.e., autoimmune encephalitis), supported by clinical red flags or abnormalities on MRI brain, EEG, or serum auto-antibody testing^3,4,6–9^, others support incorporating LP into routine testing of FEP^10^.

The lack of international consensus regarding the role of LP in the evaluation of FEP is likely driven, in part, by the absence of high-quality estimates of the prevalence of clinically relevant cerebrospinal fluid (CSF) abnormalities in this population. Existing reports of such abnormalities are derived from individual studies with limited statistical power. ^3,8^

Although there have been systematic reviews and meta-analyses^11,12^ that have examined the proportion of patients with psychosis who have CSF alterations, usefulness in general clinical practice is limited by the fact that they focused on cohorts in which secondary causes had already been exhaustively excluded (e.g., Pavel et al., Campana et al., Warren et al.). Evaluating the clinical utility of CSF analysis for FEP requires inclusion not only of these types of patients, but also of patients with undifferentiated FEP early in presentation, as well as those that might have some symptoms or signs concerning for secondary causes — these groups represent the types of cases seen at primary care offices, emergency rooms, general medical hospitals, not just the types of cases seen at specialized primary psychosis treatment programs after they have had a comprehensive workup to rule out secondary causes.

In this study, we sought to generate the first comprehensive meta-analytic estimate of the prevalence of clinically relevant CSF abnormalities in FEP. We also sought to explore how the prevalence of CSF abnormalities varied based on the *a priori* suspicion of an underlying secondary cause by grouping studies according to whether they were based on: (1) unselected FEP samples recruited solely on the basis of first-episode psychosis, regardless of whether there was a clinical suspicion for a secondary cause, (2) FEP samples with a high-clinical suspicion for a secondary cause and (3) FEP samples with low-clinical suspicion for a secondary cause.

## Methods

We performed a systematic review and meta-analysis according to the Preferred Reporting Items for Systematic Reviews and Meta-analyses (PRISMA) and Meta-Analysis of Observational Studies in Epidemiology (MOOSE) guidelines^13–15^. The study was prospectively registered on PROSPERO (CRD42024577715). A comprehensive search of Ovid MEDLINE, Embase, Web of Science, and PsychInfo was conducted up to October 2024. References of included articles and reviews were also searched. After the automated removal of duplicate papers, a title and abstract screen was performed by two independent raters, followed by a full-text review. For further details on the search strategy, eligibility criteria, and method for data extraction, quality assessment, heterogeneity assessment, and sensitivity analyses, see eMethods.

### Outcomes

Only CSF tests that were clinically available (as defined by inclusion in either of the catalogs of 2 major clinical laboratories, the Mayo Clinic Laboratories and Associated Regional and University Pathologists (ARUP) Laboratories). For consistency across studies, a common threshold was used to define an abnormal test result based on established, aligned cutoff values used by these two laboratories. For example, studies varied in the cutoff used for pleocytosis; we used >5 WBCs, which is the cut-off established by Mayo Clinic laboratories. Studies that only reported research-grade assays and continuous variables (that did not enable prevalence-estimates) were excluded. We performed meta-analytic estimates for any CSF tests that were reported by ≥3 studies. A subset CSF abnormalities were highlighted as clinically-relevant based on expert clinical opinion^4^: pleocytosis, OCB2, and antibodies associated with autoimmune encephalitis (anti-NMDAR antibodies, GAD65 antibodies, LGI1 antibodies, AMPAR antibodies, CASPR antibodies, GABAR antibodies, and Yo antibodies).

### Statistical Analysis

The overall prevalence of CSF abnormalities amongst FEP patients was calculated within each study along with a 95% confidence interval. Meta-analytic estimates were calculated for abnormalities that were reported in 3 or more studies. A random-effects model using the DerSimonian and Laird method was performed^16^. We opted to use a random-effects model as we anticipated methodological heterogeneity between studies. All analyses were performed using R 4.4.1 with meta-analyses performed using meta, metafor and metainf. Number needed to assess (NNA) for each of the abnormalities as defined by 1/proportion. We aimed to calculate relative risk of any CSF abnormalities that were reported in 3 or more studies that also had healthy controls. Additional details regarding statistical methods are included in eMethods.

### Subgroup Analyses

Where there were sufficient data (i.e. 6 or more studies)^17^, we performed subgroup analysis of 3 cohort types: (1) Unselected FEP cohorts that included all individuals with first-episode psychosis, irrespective of clinical suspicion for a secondary cause, (2) cohorts that selected for patients with a high-clinical suspicion for a secondary cause and (3) studies that specifically excluded patients with high-clinical suspicion for secondary causes. In addition, for covariates that had data from at least 6 studies, meta-regression was used to examine their effects; only age and publication year met these criteria^17^.

## Results

### Search Results and Study Selection

The search strategy resulted in 2,476 publications. After the removal of duplicates, screening of title and abstracts, 257 papers were reviewed in full. eFigure 1 shows the PRISMA flow chart and additional details are in eMethods.

### Study Characteristics

There were 38 eligible papers^4,6,11–50^; reporting findings from 32 cohorts identified. See eTable 1 in Supplement 1 for detailed study characteristics. Papers were published between 1978 and 2024, reporting a total sample of 3,680 FEP patients. One paper was excluded due to high risk of bias^42^ resulting in 37 papers being included with 3,330 unique patients. Only 3 papers reported healthy control comparison groups^19,34,52^. Studies were conducted in the following regions: Europe (n=28), North America (n=6), and Asia (n=4). While all patients within these studies received an LP, studies selected their patient population in different ways.19 papers had cohorts that excluded patients with suspicion for secondary causes for FEP (e.g. suspected autoimmune encephalitis), 5 papers selected for patients with a high suspicion for secondary causes, 2 papers had cohorts that were separable into a discrete high-suspicion group and discrete low-suspicion group, and the remaining 12 papers were based on an undifferentiated sample. Studies were not uniform in how they determined high or low suspicion for secondary psychoses with different clinical or ancillary testing used (see eTable 3 for details of individual methods).

### Participant Characteristics

FEP sample sizes ranged from 13 to 332. Mean age ranged from 20-49 years. The median proportion of female was 42% (IQR 20.1 to 57.5). Duration of untreated psychosis ranged from 4 to 103.8 weeks. Practice settings included: inpatient psychiatric unit at an academic medical center (18), inpatient psychiatry and neurology unit within a general hospital, research ward or unspecified setting (8), mixed setting spanning inpatient units, a neurological center, and outpatient settings (3), acute (emergency) psychiatry (2), and finally, academic medical center (1).

### Characteristics of CSF analysis

Assays reported in 3 or more studies were CSF IgG index, protein, OCBs Type 2, Type 3, Type 4, and Type 5, Cell Count (both studies using the more stringent WBC >5, and studies using WBC≥5), anti-NMDAR antibodies, GAD65 antibodies, LGI1 antibodies, AMPAR antibodies, CASPR antibodies, GABAR antibodies, and Yo antibodies.

### Quality Assessment and Risk of Bias

Twenty-four cohorts were low risk of bias, 7 were medium risk of bias, and 1 was high risk of bias^42^ (and was therefore excluded).

### Prevalence of clinically relevant CSF abnormalities

Pleocytosis (defined as > 5WBCs) (Figure 1a) had a prevalence of 3.2% (95% CI 2.1-4.4, n=1087), a number needed to assess (NNA) of 31, and an I^2^ of 0.0%. OCBs restricted to the CSF (Type 2)^58^ (Figure 1b) had a prevalence of 7.1% (95% CI 3.6-11.7, n=1400), a NNA of 14, and an I^2^ of 84.8%. Anti-NMDAR antibodies (Figure 1c) had a prevalence of 2.3% (95% CI 0.1-6.4, n=1279), a NNA of 43, and an I^2^ of 87.1%. Yo antibodies (eFigure 2a) had a prevalence of 0.4% (95% CI 0 to 1.7, I^2^ 14.3%, n=367). GAD65, LGI1 antibodies, AMPAR, CASPR, and GABAR antibodies (eFigure 2) all had a prevalence of 0% (95% CI 0 to 0.3, I^2^ 0.0%, n=507 [GAD65], n=359 [LGI1], n=395 [AMPAR and GABABR], n = 365 [CASPR]) in the CSF.

**Figure 1.**
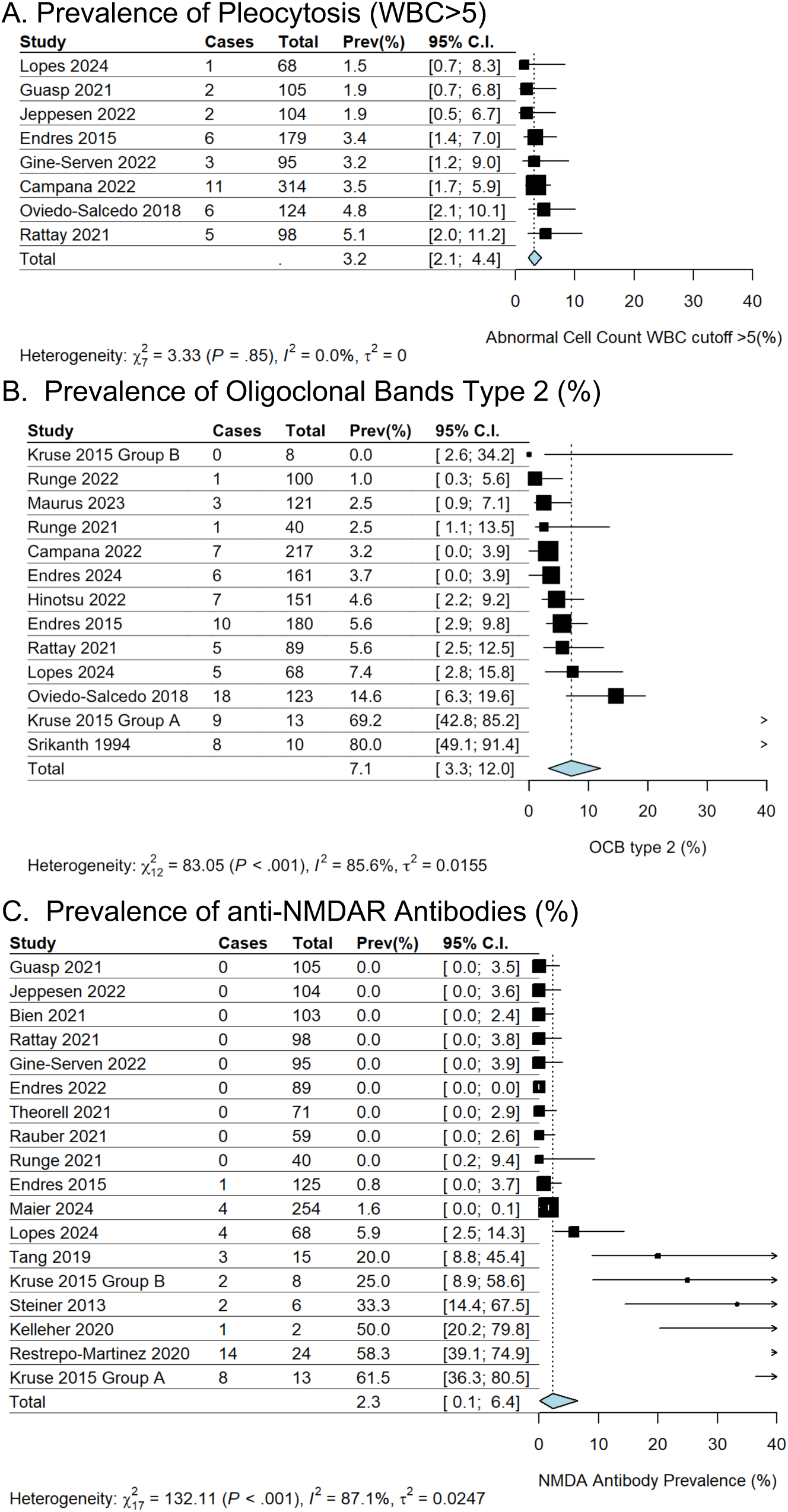
Forest plots of clinically relevant CSF abnormalities in first-episode psychosis. The size of each box is proportional to the weight of the study in relation to the pooled prevalence estimate (as calculated by a random effects model in the final row of the figure). Columns: Cases – number of FEP patients with that particular CSF abnormality, Total – total number of FEP patients tested, Prev (%) – raw prevalence of abnormality in that study, C.I. – Confidence Interval. Kruse 2015 Group A – patients who had serum positivity for anti-NMDAR antibodies. Kruse 2015 Group B-all patients who had serum studies regardless of NMDAR positivity

### Prevalences of other CSF abnormalities (eFigure 2)

Elevated CSF IgG index had a prevalence of 5.4% (95% CI of 1.1 – 11.6, I^2^ 78.6%, n=526). Type 3 OCBs had a prevalence of 6.1% (95% CI 3.3 – 9.5, I^2^ 53.0%, n=546), Type 4 OCBs had a prevalence of 9.6% (95% CI 3.1 – 19.0, I^2^ 92.4%, n=717), and Type 5 OCBs had a prevalence of 0% (95% CI 0 - 0.5, I^2^ 0.0%, n=338). Prevalence of elevated albumin was 19.5% (95% CI 14.7 - 24.7, I^2^ 58.5%, n=782) and elevated total protein was 18.7% (95% CI 9.7 to 29.5, I^2^ 93.5%, n = 1,194).

### Relative Risk of CSF Abnormalities

Relative risk was not calculated as there were ≤ 3 study cohorts with healthy control groups reporting values for any given test^19,34,52^.

### Subgroup Analyses

Subgroup analyses are shown in Figure 2. The prevalence of pleocytosis (WBC>5) was 3.1% (95% CI 1.9-4.6, k=5, I^2^ 0%, n=742) amongst studies that excluded cases with clinical suspicion of secondary causes. Subgroup estimates could not be made for the undifferentiated or high clinical suspicion subgroups because each had less than 3 studies.

**Figure 2.**
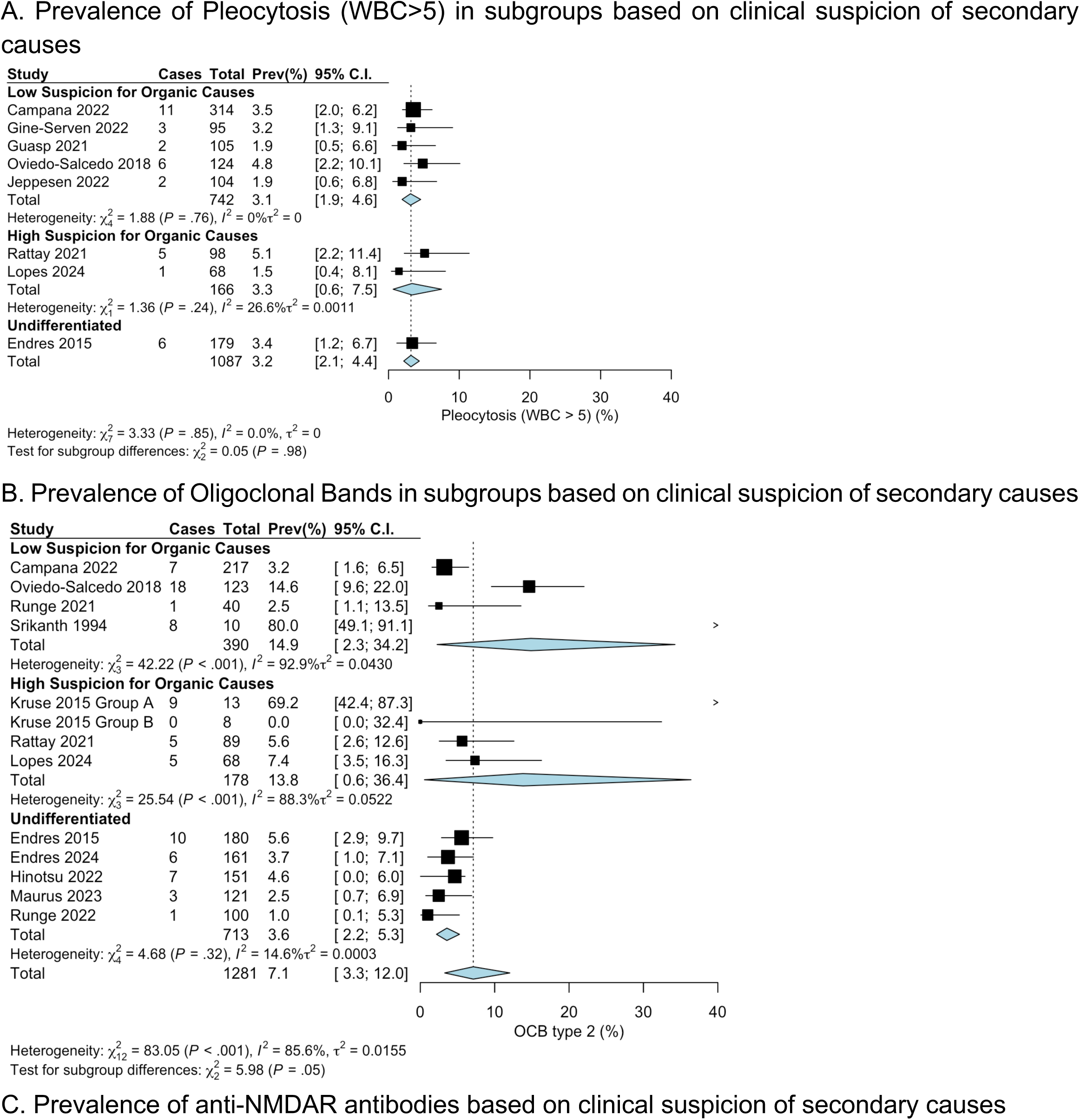
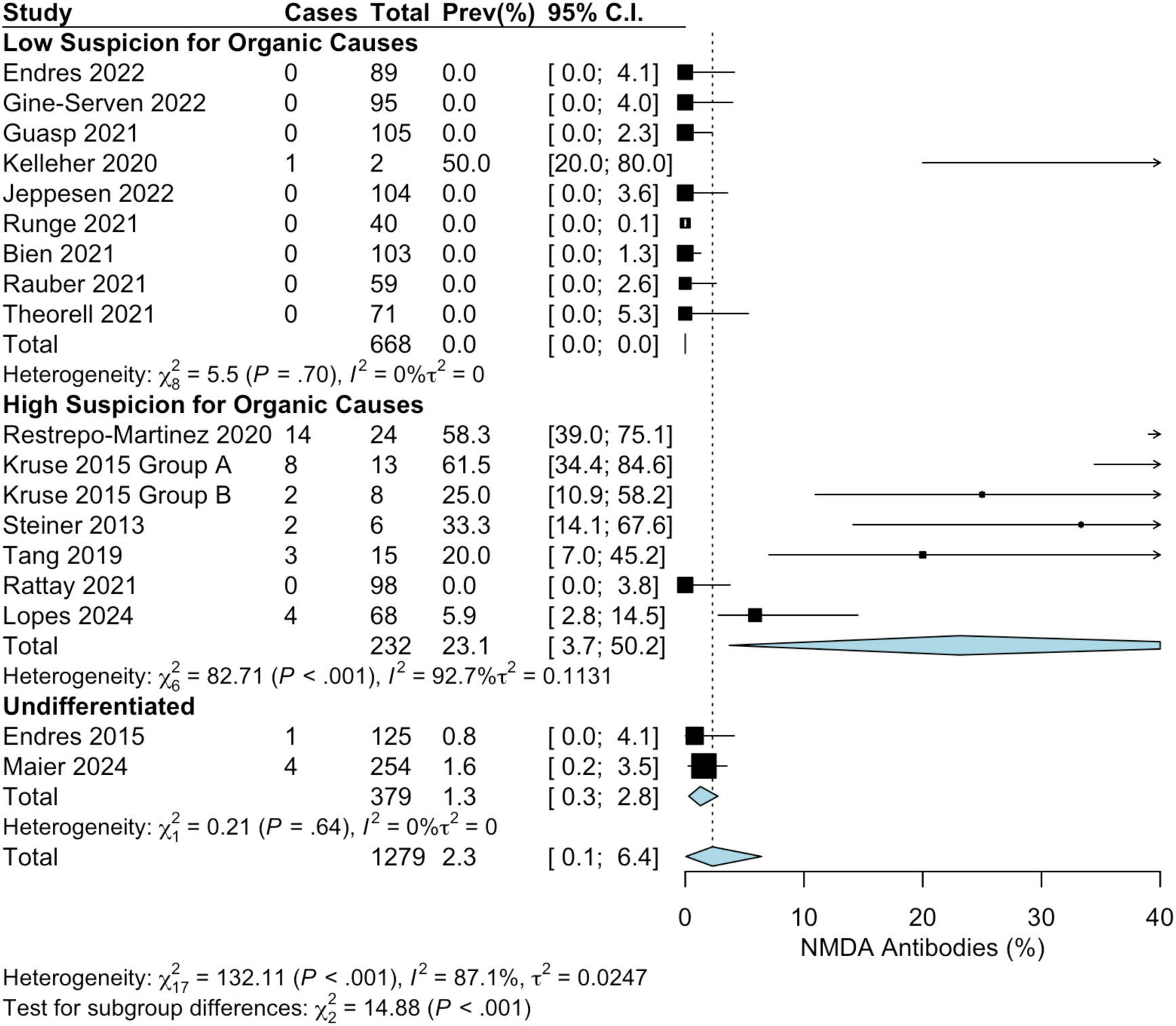
Forest plots of subgroup analysis of clinically relevant lumbar puncture abnormalities in First-Episode Psychosis, based on clinical suspicion of secondary causes. The size of each box is proportional to the weight of the study in relation to the pooled prevalence estimate (as calculated by a random effects model in the final row of the figure). Columns: Cases – number of FEP patients with that particular CSF abnormality, Total – total number of FEP patients tested, Prev (%) – raw prevalence of abnormality in that study, C.I. – Confidence Interval. Kruse 2015 Group A – patients who had serum positivity for anti-NMDAR antibodies. Kruse 2015 Group B-all patients who had serum studies regardless of NMDAR positivity.

The prevalence of OCB2 was 14.9% (95% CI 2.3-34.2, k=4, I^2^ 92.9%, n=390) amongst studies that excluded cases with clinical suspicion of secondary causes. Prevalence was 3.6% (95% CI 2.2-5.3, k=5, I^2^ 14.6%, n=713) amongst studies that did not include or exclude cases based on clinical suspicion of secondary causes. Prevalence was 13.8% (95% CI 0.6-36.4, k=4, I^2^ 88.3%, n=178) for the high clinical suspicion subgroup.

The prevalence of anti-NMDAR antibodies was 0% amongst studies that excluded cases with clinical suspicion of secondary causes (95% CI 0.0-0.0%, I^2^ 0%, n=668), was 1.3% (95% CI 0.3 to 2.8, k=3, I^2^=0%, n=452) amongst studies that did not include or exclude cases based on clinical suspicion of secondary causes, and was 23.1% (95% CI 3.7-50.2, k=7, I^2^=92.7%, n=230) in the subgroup with high clinical suspicion of secondary causes.

### Influence of Potential Effect-Modifiers and Publication Effects

Meta-regression found year of publication did not have a significant effect on pleocytosis (p = 0.46) but was associated with reduced OCB2 (p < 0.0001) and anti-NMDAR antibodies (p < 0.005). No statistically significant associations were identified for age for pleocytosis (p = 0.82), OCB2 (p = 0.27), or anti-NMDAR antibodies (p = 0.93). Leave-one-out analyses showed general stability of pooled prevalence and heterogeneity estimates, suggesting that the overall results are not driven by any individual dataset (Fig. 3A). Funnel plots (Fig. 3B) showed no significant asymmetry by Egger’s test for pleocytosis (p = 0.71) but did show significant asymmetry for OCB2 and anti-NMDAR antibodies (p < 0.0001), suggesting the latter may have small-study effects or publication bias (e.g., small studies tended to have higher prevalence of anti-NMDAR antibodies), or that studies included populations with different a priori risks of OCB2 or anti-NMDAR antibodies

**Figure 3.**
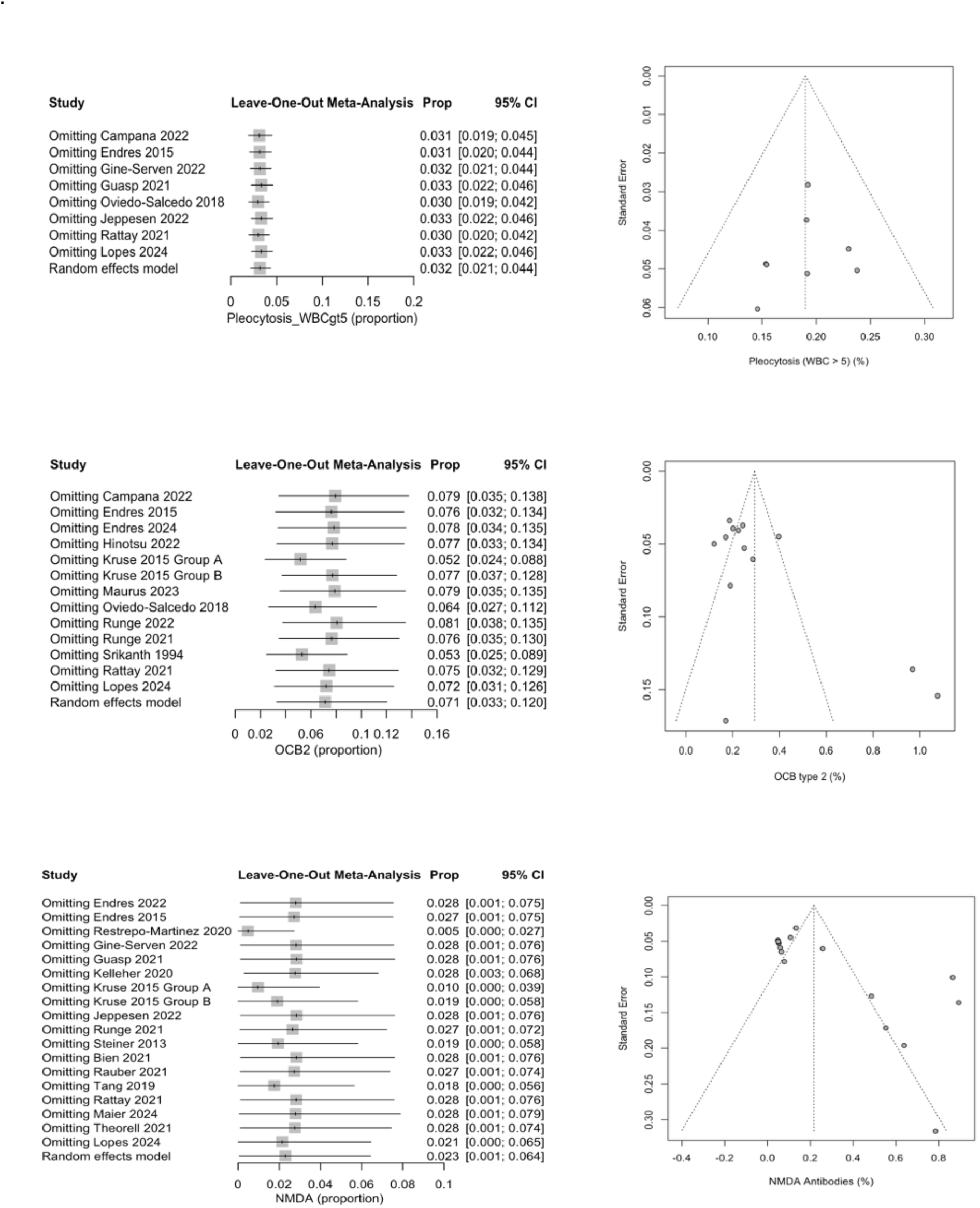
Publication Effects. A) Leave one out analyses show influences of individual studies B) Funnel plots show potential for small study effects, publication bias, or differences in baseline characteristics in studies.

## Discussion

We report the first comprehensive meta-analytic prevalence-estimates of clinically relevant cerebrospinal fluid (CSF) abnormalities in patients with first-episode psychosis (FEP).

The most common finding was OCB2 (7.1%), followed by pleocytosis (3.2%), anti-NMDAR antibodies (2.3%) and other anti-neuronal antibodies (0.4%). The presence of pleocytosis, CSF auto-antibodies^4,59^, or OBC2 may alter prognosis or management in the appropriate clinical context, including consideration of further ancillary testing or immunomodulatory treatments that would not form part of the normal treatment pathway of a primary psychotic disorder^60,61^. At 7.1%, the prevalence of OCB2 in FEP is comparable to the prevalence of a clinically relevant abnormality on MRI brain based on recent meta-analytic estimates^62^. As another benchmark for reference, CT head has an estimated diagnostic yield close to 0% in the workup of FEP^63^. The prevalence of OCBs in cryptogenic epilepsies (8%) and acute-Covid related neurological symptoms (2%), for which LP are sometimes pursued, suggest that lumbar puncture should be considered as part of the clinical workup in FEP.

The current initial workup for FEP varies between hospital systems but often includes serum laboratory testing, with the addition of neuroimaging and EEG^64^ in some centers. Though lumbar puncture is not often included as a part of this initial workup, outside of a few hospital systems in Germany (as per the Freiburg diagnostic protocol for FEP^65^), it is often recommended in cases whether there is a heightened suspicion of autoimmune encephalitis based upon clinical history, exam, and/or other testing results^3^. Subgroup analyses in our analyses suggested that including, or excluding patients with high clinical suspicion for secondary psychosis makes a substantial difference to the detection of anti-neuronal antibodies. Prevalence of anti-NMDAR antibodies was an order of magnitude higher in the high-clinical suspicion group (23.1%) compared to samples with undifferentiated FEP (1.3%). In contrast they were undetectable in samples which excluded high-suspicion individuals. These observations suggest that thorough clinical assessment is important, and LP might be viewed as a critical diagnostic tool in people with FEP and high suspicion for secondary causes; an important technical caveat being, however, that the methods studies used to define high suspicion differed (see supplementary materials).

The prevalence of OCBs was marginally higher in the high-clinical suspicion group (13.8%) than in an undifferentiated FEP group (3.6%), studies that excluded patients considered high risk of a secondary psychosis did not have lower prevalence (14.9%);This suggests that the clinical protocols for determining low suspicion groups did not effectively exclude some clinically relevant CSF abnormalities.

Our results also raise the question of what should be the clinical role of LP in FEP. Although the prevalence of CSF abnormalities in FEP provides critical information, there are other important factors to consider prior to determining the clinical place of LP in FEP such as safety, acceptability, cost, and accessibility. LPs are generally safe procedures, even amongst other vulnerable populations such as patients with Alzheimer’s dementia^67^. The most common iatrogenic risk is post–LP headache^68,69^ and the risk of more serious complications is low at 0.20%^70^. A recent meta-analysis found that post-LP headache prevalence can be reduced to 4% with the incorporation of novel methods (i.e., atraumatic needle) as compared to 11% with the conventional method (i.e., traumatic needle)^71^. Acceptance rates of LP amongst patients with psychosis have been estimated at 66%, with acceptance influenced by factors such as informative patient education, perceived risk of invasiveness, and perceived necessity^72,73^ Formal cost-benefit analyses of LP in FEP are lacking, though there has been one USA-based study on cost-effectiveness of serum testing for anti-NMDAR antibodies in all cases of FEP to select high-risk individuals needing LP and CSF analyses for anti-NMDAR antibodies^74^. Accessibility to LP is geographically variable, and patient factors such as having MRI-incompatible implanted device(s) may preclude the accessibility of alternative imaging-guided approaches.

### Strengths and Limitations

A meta-analytic estimate transcends the limits of small individual sample size and study-design variation, thus rising to the highest category of quality by the Oxford Centre for Evidence-Based Medicine criteria (cebm.net). A meta-analysis of prevalence of blood brain barrier disruption in N=531 patients with recent onset psychosis was recently reported^75^, outside the dates of our literature search; our meta-analysis has a combined sample nearly seven times larger than this, focuses on the most clinically relevant CSF abnormalities, and has a more comprehensive inclusion of clinical abnormalities. Our study’s general strengths include its comprehensive search strategy, adherence to PRISMA and MOOSE guidelines, and rigorous assessment of heterogeneity and publication bias. Subgroup analyses allowed us to explore potential moderators and sources of heterogeneity.

However, several limitations also warrant consideration. First, most studies did not include healthy controls, preventing us from being able to explore the relative risk of CSF abnormalities in psychosis. Second, heterogeneity across studies was substantial, with I² values exceeding 80% for OCBs and anti-NMDAR antibodies, likely reflecting variability in the included patient populations, diagnostic criteria, and laboratory methods. Moreover, though our study assembled a comparatively large sample size, the low prevalence of some abnormalities still resulted in wide confidence intervals, consistent with previous meta-analyses on prevalence of anti-NMDAR antibodies in serum^76^. Our Egger plot analysis also suggested possible small-study effects, cohort differences, or publication bias of some abnormalities.

The majority of studies were from a population representative of patients with FEP, but certain studies had selected patient populations that may be influencing prevalence-estimates. For example, one study^23^ examined only COVID-19 positive patients and another study^39^, included patients with high clinical suspicion for “organic” psychosis between 1998 and 2016, but thereafter included all patients with psychosis. Notably, year of publication was associated with declining prevalence of OCBs, pleocytosis and anti-NMDAR antibodies, potentially reflecting advances in diagnostic specificity or changes in patient selection over time. Sometimes we were able to detect differences in reporting; for example, studies differed on the WBC cutoff to define pleocytosis, and we were able to harmonize the cutoff (in this case to greater than 5 WBCs). Other methodological differences in how studies conducted CSF testing were also not able to be accounted for in this study, for example, there increasing attention is being paid to the effect of different methods of anti-neuronal antibody ascertainment^77,78^ and studies included here did not use identical methods.

Importantly, as studies did not consistently report individual patient-specific CSF abnormalities, we are not able to generate a pooled prevalence-estimate of likelihood of having *any (i.e at least one)* clinically relevant abnormality. Such information would be informative in practice, for example, as an individual may have a pathogenic process that is evident by detecting abnormality in one of several possible tested components, i.e., a pathogenic neuroimmunological process may show itself in a particular individual with the presence of an autoantibody, inflammatory routine studies, or elevated oligoclonal bands, but not necessarily all of the abnormalities or multiple of the abnormalities concurrently. The conservative estimate of the prevalence of *at least one* clinically relevant CSF abnormality is 7.1%, the rate of OCB2, but the true prevalence of having *any* abnormality is likely higher (e.g. as some people might have pleocytosis but not OCB2s).

Finally, our study was limited to a binary definition of most variables as abnormal or not, and any additional information from magnitude of the abnormality was not captured. For example, for anti-CNS antibodies, many studies reported only presence or absence, though the antibody titer is often important for interpretation.

### Conclusion and perspectives

This meta-analysis found the prevalence of clinically-relevant CSF abnormalities in FEP to be at least 7.1%, and therefore comparable to the prevalence of clinically relevant imaging abnormalities via brain MRI. The prevalence of clinically-relevant CSF abnormalities was higher amongst patients with a high clinical suspicion for secondary causes. The findings underscore the need for continued consideration of LP as a diagnostic approach for the evaluation of FEP, and this meta-analytic estimate of CSF abnormality prevalence provides current state-of-the-art evidence to guide the decision whether to pursue LP. Having established the prevalence of clinically relevant CSF abnormalities in FEP, further evidence is needed to determine the clinical utility of lumbar puncture in this patient group, however. This includes research on its diagnostic impact, treatment implications, effects on clinical outcomes, acceptability to patients and clinicians, and cost-effectiveness. Moreover, further studies are needed to compare effectiveness of different diagnostic algorithms for secondary psychoses and to report the clinical outcomes of patients with CSF abnormalities (especially those without an identified, clinically well-defined auto-antibody), including response to targeted treatments.

## Data Availability

All data produced in the current study are available upon reasonable request to the authors.

## Acknowledgements

Thank you to Tim W. Rattay, MD and Dirk Wildgruber, MD for generously providing further detail of the FEP patients with CSF results from Rattay et. al, 2021, to enable those patients to be included in this meta-analysis. MLB receives royalties from Oxford University Press for the sale of his book, “The Neuroethics of Biomarkers: What the Development of Bioprediction Means for Moral Responsibility, Justice, and the Nature of Mental Disorder.” The authors have no conflicts of interest.

## Supplement

### eMethods

#### Search Strategy

The search was conducted in a stepwise fashion: (1) electronic databases were searched from inception until October 2024, and (2) the references of eligible articles and reviews were manually reviewed. A title and abstract screen was performed prior to full article review to confirm eligibility. Databases included were Ovid, MEDLINE, Embase, PsychINFO Web of Science using the search terms that were developed in consultation with a research librarian (AF). Two researchers (A.C., L.D., A.P., M.B) independently performed the title and abstract screen and full text review in parallel. Articles with consensus approval between both raters were included. Discrepancies were resolved by an independent rater (GB).

#### Search Terms

##### OVID Medline

Schizophrenia/di OR Psychotic Disorders/di OR (psychotic* OR psychosis or schizophren*).ti,ab,kw,kf

AND

exp cerebrospinal fluid/ OR exp spinal puncture/ OR (cerebrospinal fluid OR CSF OR spinal puncture* OR lumbar puncture*).ti,ab,kw,kf

AND

Diagnosis/ OR (diagnos*).ti,ab,kw,kf

##### Embase

’schizophrenia’/mj OR ’psychosis’/mj OR (psychotic* OR psychosis or schizophren*):ti,ab,kw

AND

’cerebrospinal fluid’/exp OR ’lumbar puncture’/exp OR (’cerebrospinal fluid’ OR CSF OR ’spinal puncture*’ OR ’lumbar puncture*’):ti,ab,kw

AND

’diagnosis’/mj OR (diagnos*):ti,ab,kw

##### Web of Science

TI=(psychotic* OR psychosis or schizophren*) OR AB=(psychotic* OR psychosis or schizophren*) OR AK=(psychotic* OR psychosis or schizophren*)

AND

TI=(“cerebrospinal fluid” OR CSF OR “spinal puncture*” OR “lumbar puncture*”) OR AB=(“cerebrospinal fluid” OR CSF OR “spinal puncture*” OR “lumbar puncture*”) OR AK=(“cerebrospinal fluid” OR CSF OR “spinal puncture*” OR “lumbar puncture*”)

AND

TI=(diagnos*) OR AB=(diagnos*) OR AK=(diagnos*)

##### PsychInfo

(DE “Schizophrenia” OR DE “Acute Schizophrenia” OR DE “Catatonic Schizophrenia” OR DE “Paranoid Schizophrenia” OR DE “Process Schizophrenia” OR DE “Schizoaffective Disorder” OR DE “Schizophrenia (Disorganized Type)” OR DE “Schizophreniform Disorder” OR DE “Undifferentiated Schizophrenia”) OR TI (psychotic* OR psychosis OR schizophren*) OR AB (psychotic* OR psychosis or schizophren*)

AND

DE “Cerebrospinal Fluid” OR TI (“cerebrospinal fluid” OR CSF OR “spinal puncture*” OR “lumbar puncture*”) OR AB (“cerebrospinal fluid” OR CSF OR “spinal puncture*” OR “lumbar puncture*”)

AND

MM “Diagnosis” OR TI (diagnos*) OR AB (diagnos*)

#### Eligibility Criteria

Eligibility criteria were FEP patients (including patients with psychosis possibly due to a general medical condition, primary psychiatric disorder, and unspecified psychosis) for whom clinically available CSF testing had been reported to allow for the calculation of prevalence-estimates. Studies were included if they performed one or more clinically available CSF analyses, defined as being in the laboratory manual of Mayo Clinic or ARUP. Where there were overlapping samples, the study with the larger sample was utilized. There were no age or language restrictions. Excluded from the analysis were studies with insufficient data to permit calculation of prevalence-estimates, post-mortem studies, case reports or case series.

#### Data Extraction and Encoding

Data extraction was independently performed in Covidence systematic review software (Veritas Health Innovation, Melbourne, Australia. Available at www.covidence.org) by four researchers (AC, MB, LD, AP) using a piloted standardized form with a fourth researcher (GB) resolving any discrepancies. For each study, information on the study characteristics, sample, and CSF handling processes was also extracted. Where reported, healthy control data was also extracted to calculate risk ratios. For categorizing of subgroups and modeling purposes, certain assumptions were made and the following *ad hoc* criteria were applied for borderline cases: 1) if a study only excluded patients with known dementia (but not other causes of psychosis), they were coded as an undifferentiated sample, 2) if a cohort study changed inclusion criteria mid-study from including only cases with clinical high-suspicion for secondary cause to including all FEP (i.e. undifferentiated FEP), they were coded as undifferentiated FEP provided >80% of the cohort was collected during the period where LP was performed for all FEP. If not, they were coded as high-suspicion subgroups. This grouping was done *post hoc*.

In our review of studies, we found that there were several from one research group that had nested, partially overlapping samples: Endres 2015, 2017, 2020, 2021, 2022, and 2024. All of these studies had patient samples that partially overlapped with Endres 2020, and each study had some overlap with the closest chronological study, but Endres 2015 appeared to have no sample overlap with 2022 and 2024. As such we chose to include Endres 2015 and Enders 2022+2024 (as a merged cohort extracting the largest sample for each CSF test), as this resulted in the largest non-overlapping patient cohort from the overlapping group.. Similarly, Campana 2022 and 2023 were found to have 100% overlap and as such, the larger n amongst the two publications was chosen for any given prevalence estimate.

There were some studies where a substantial portion of patients were FEP but the report did not enable the disaggregation of those with FEP and those with persistent psychosis, Rattay 2021, Runge 2021 and Runge 2022. Rattay was able to be contacted to disaggregate the results. In the case of Runge 2021 and 2022, the studys’ risk of bias rating was downgraded as it only partially met the sample selection criteria; since the final risk of bias was not “high risk”, these studies were ultimately included.

#### Quality Assessment

A 10-item risk of bias assessment tool was utilized (Hoy et al. 2012). Individual papers were categorized based on a summative score and categorized as high (0-3), moderate (4-6), or low (7-10) risk of bias^79^. If multiple papers shared a cohort but diverged in quality assessment, the average quality assessment score was used for categorization. Those deemed as having high risk of bias were excluded.

#### Heterogeneity Assessment

The I^2^ index was used to assess heterogeneity. Low degree of heterogeneity was defined as an I^2^ of <25%, moderate heterogeneity was 25 to 75%, a high degree heterogeneity was defined as 75 to 100%. A Baujat plot was also generated to assess heterogeneity and influence.

#### Sensitivity Analyses

Sensitivity analyses were performed to explore the robustness of findings, including the effects of studies with a moderate risk of bias. Influencer diagnostics were performed based on leave one out analyses to identify studies with excessive influence on the pooled prevalence-estimates or heterogeneity. Risk of publication bias was assessed via visual inspection (for asymmetry) of Egger plots^80^ for main outcomes.

#### Use of AI or LLM

Leave-one-out sensitivity analysis were conducted with the code from Blackman et al. 2023^81^ that is publicly available on github. However, due to changes in the meta package in R from the version in use in Blackman et al. 2023 to the version used for the analyses in this manuscript, the code required debugging in order to be compatible with the new version of the meta package. Chat GPT 5.0 (openAI) was used to help troubleshoot these errors. AI was not used for the generation of any of the images, plots or original analyses, or elsewhere in the manuscript.

## eFigures

**eFigure1.**
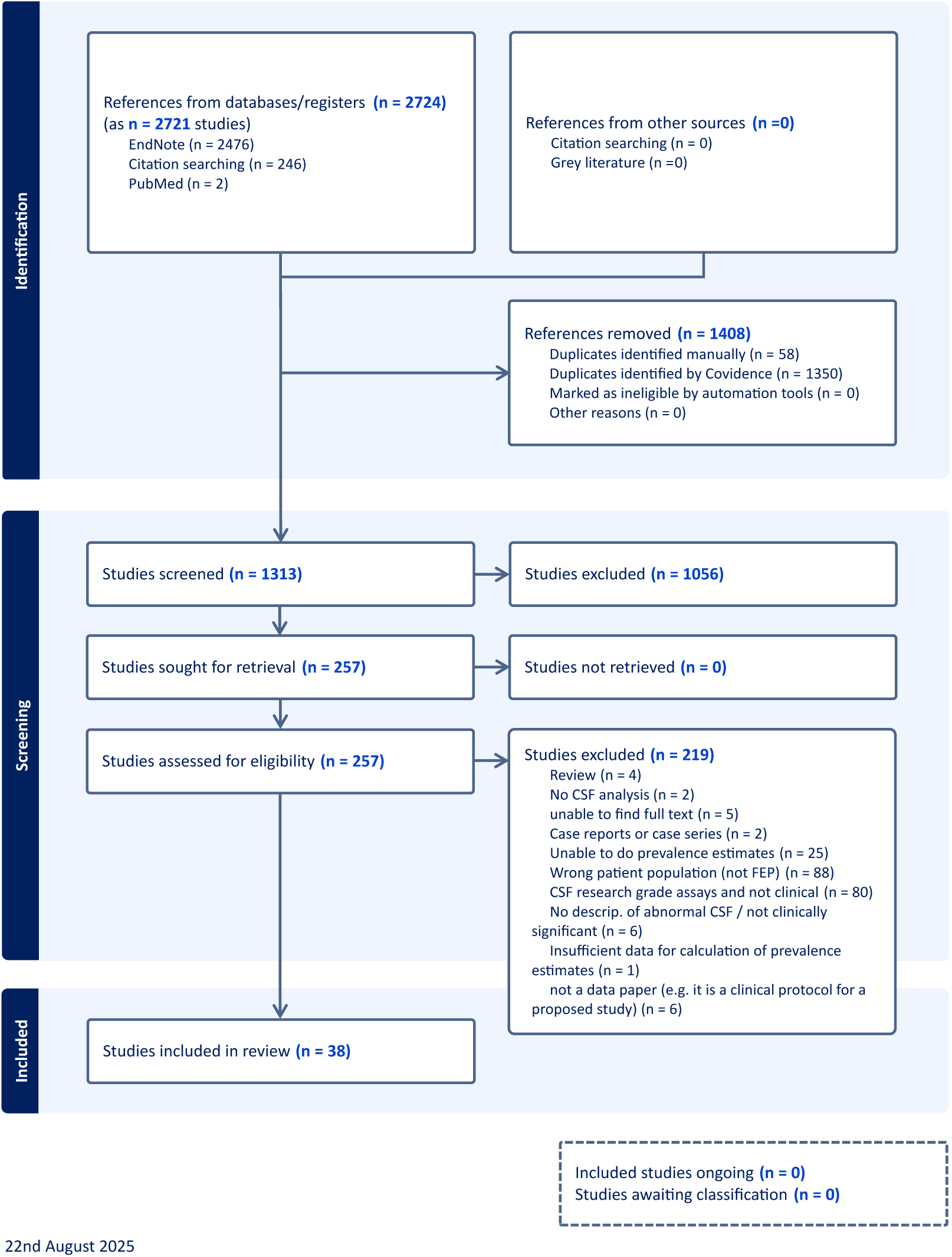
PRISMA flow diagram

**eFigure2.**
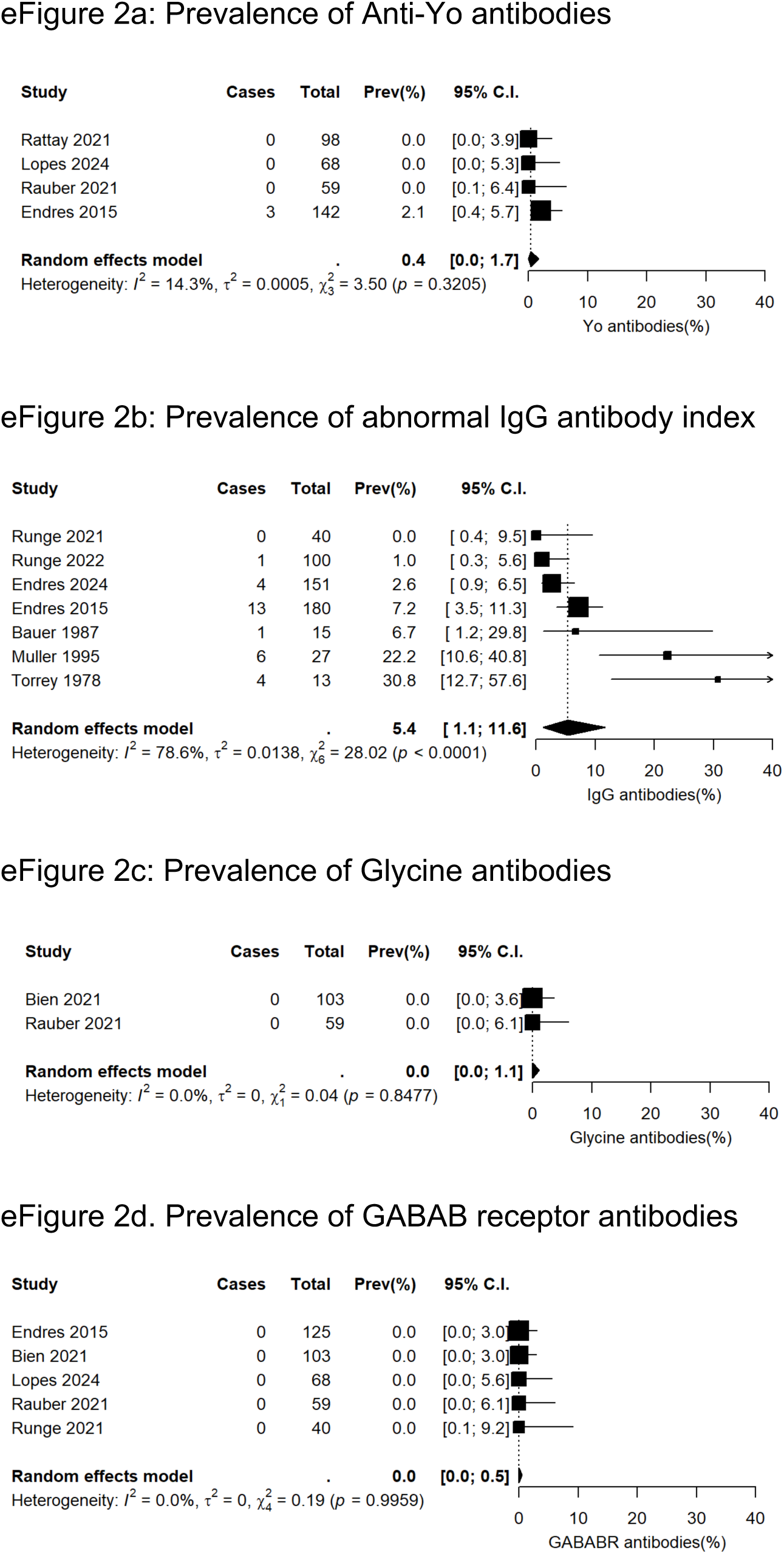
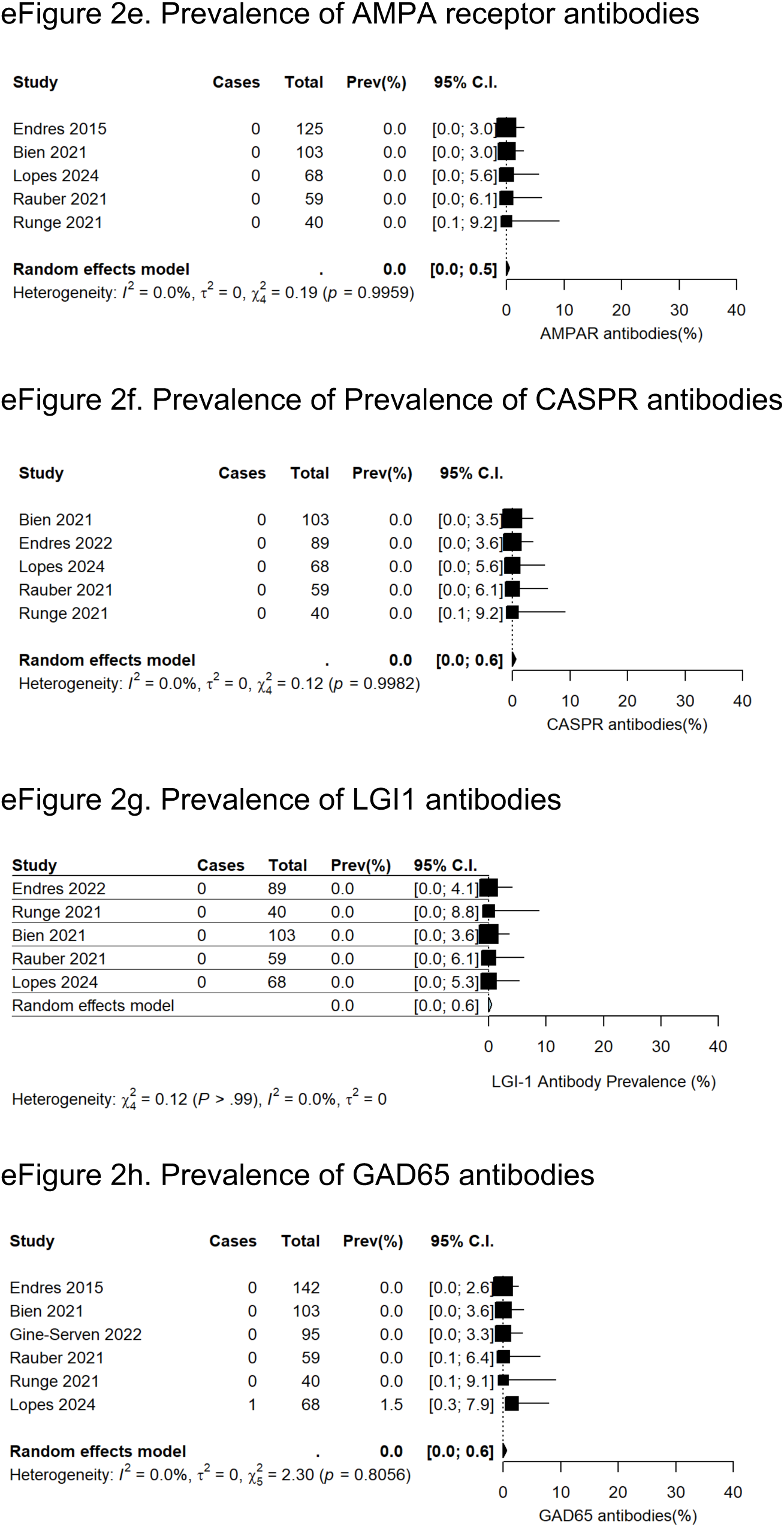
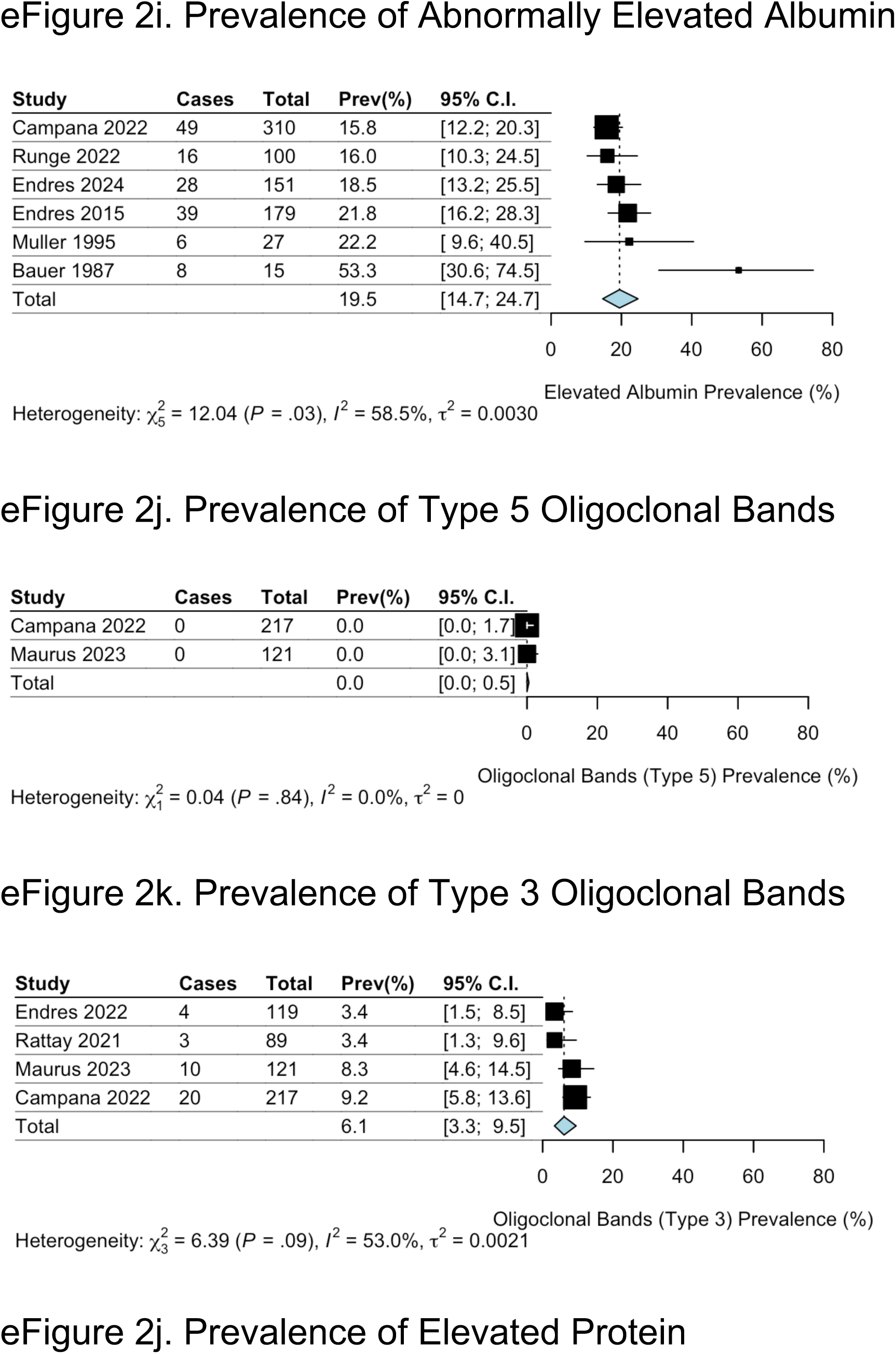
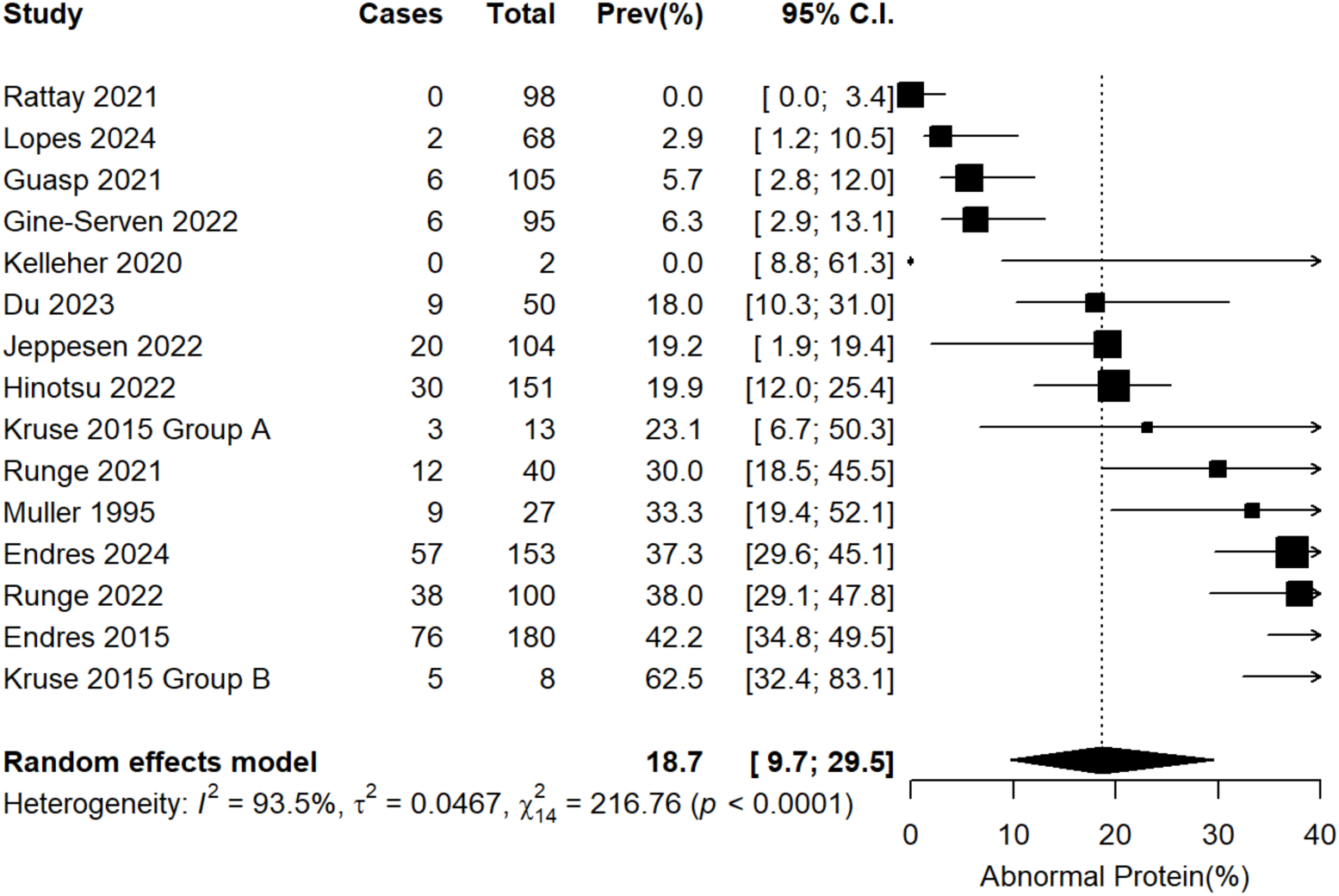
Prevalence-estimates of other CSF abnormalities

**eFigure 3.**
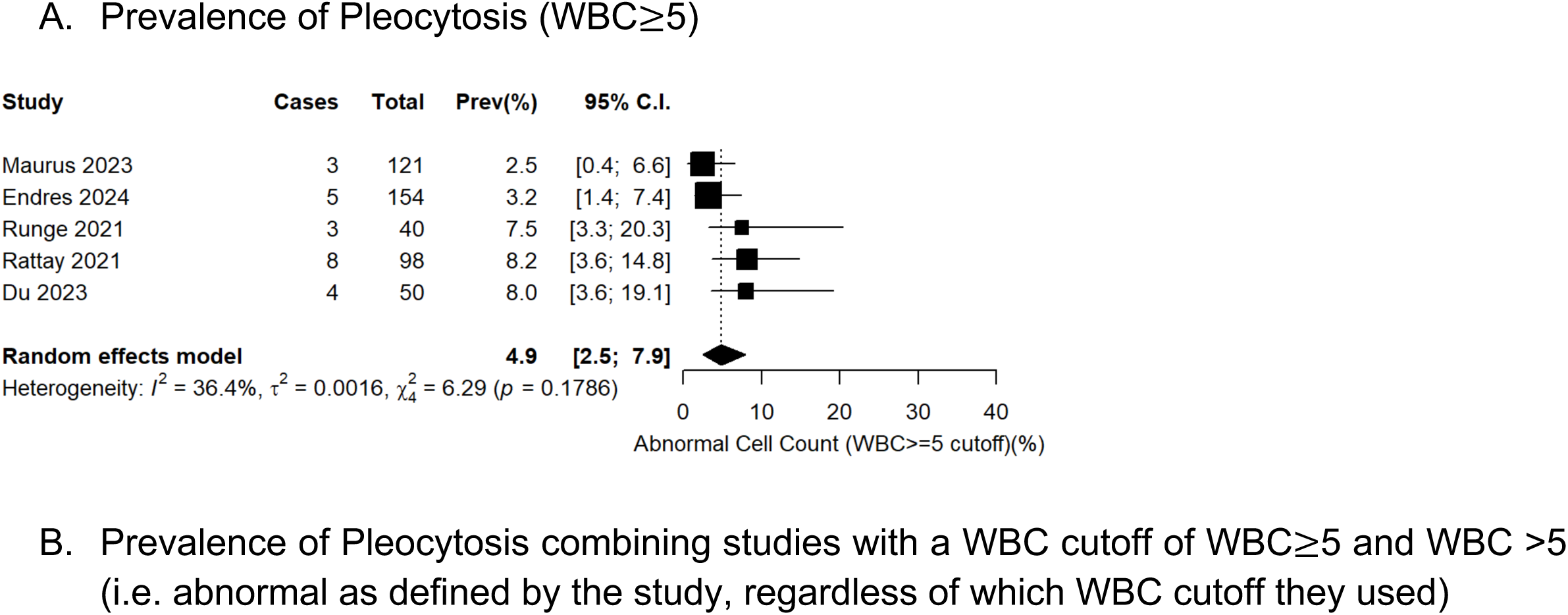
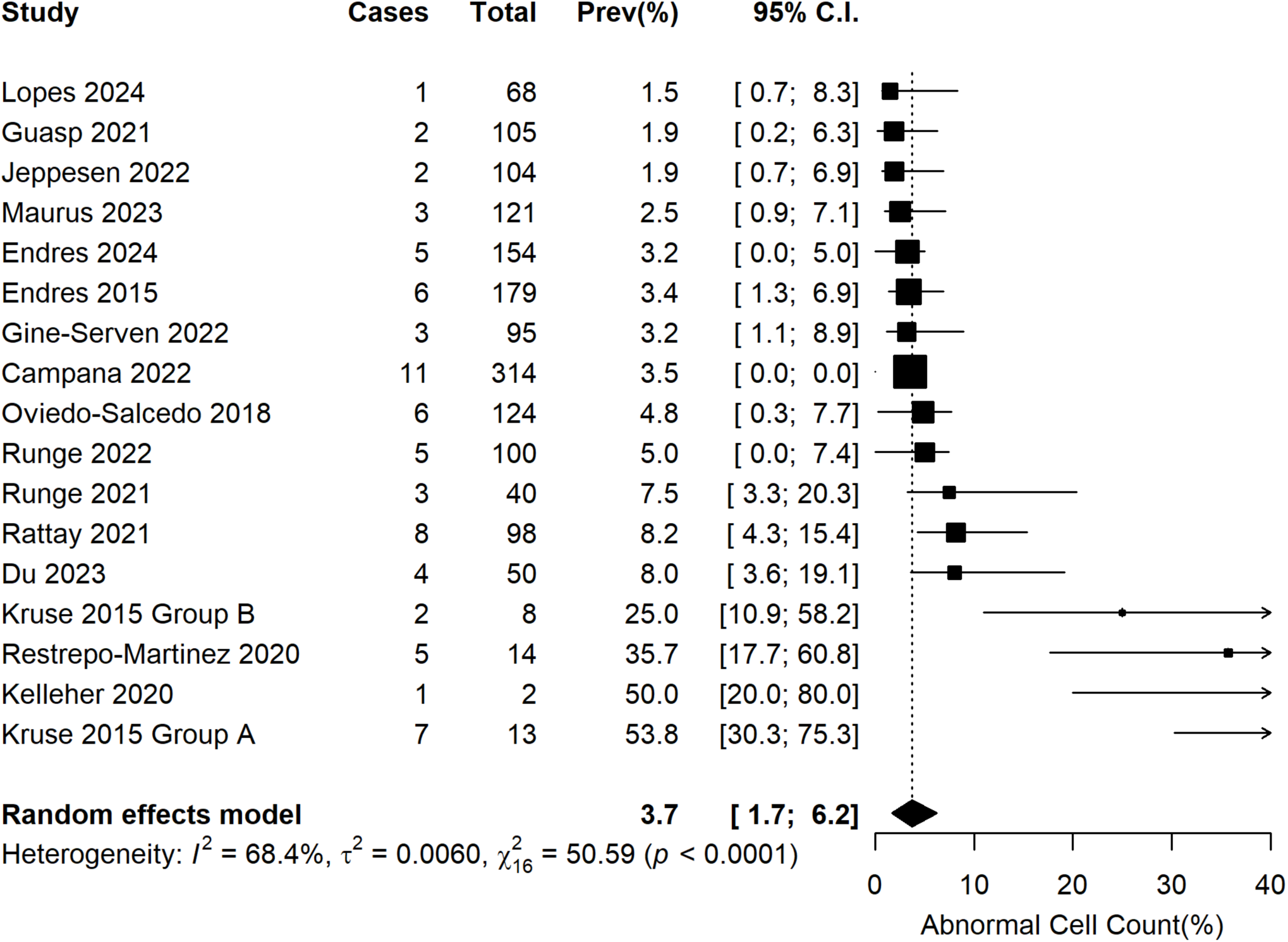
Prevalence of Pleocytosis amongst studies with other WBC cutoff values

**eFigure 4.**
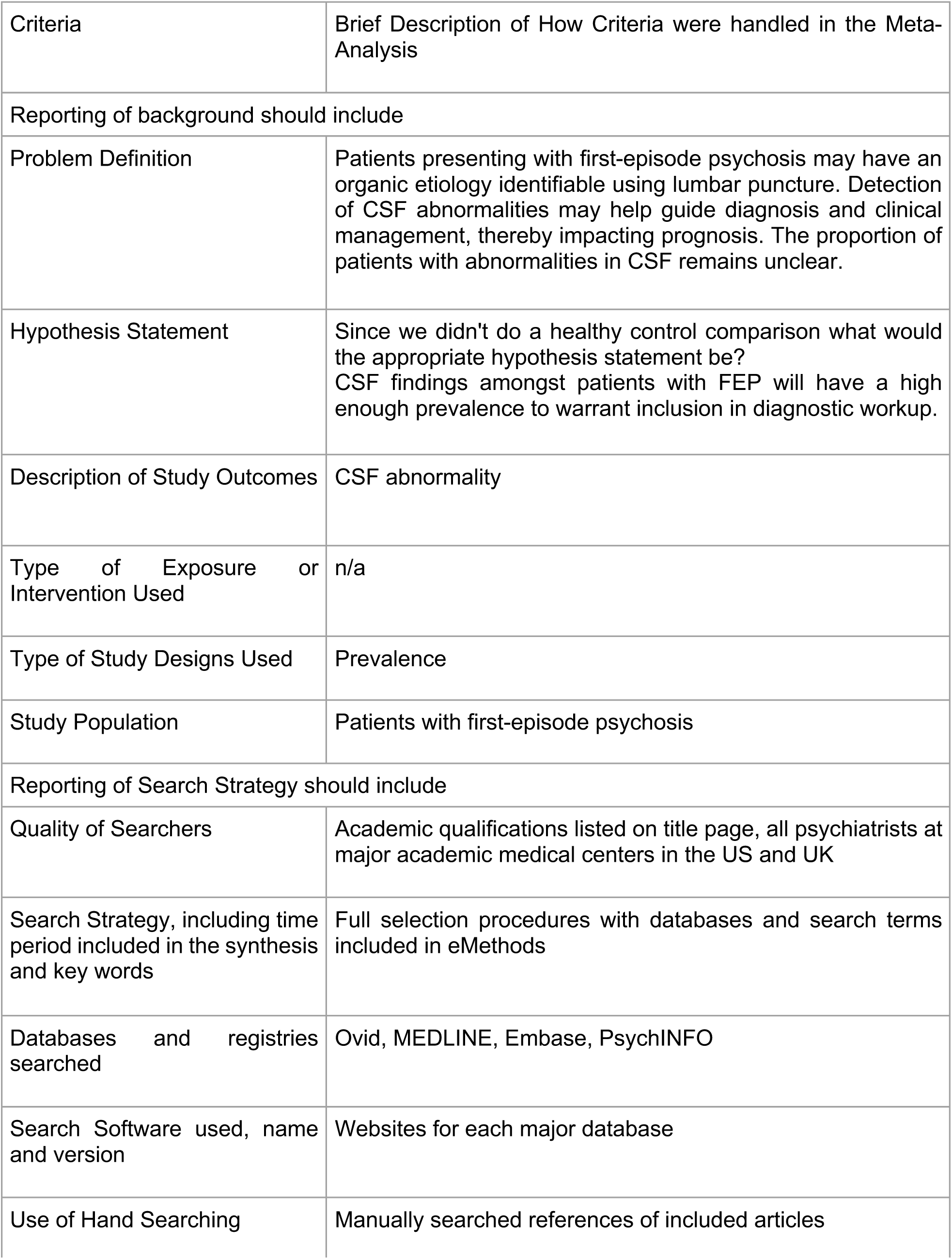
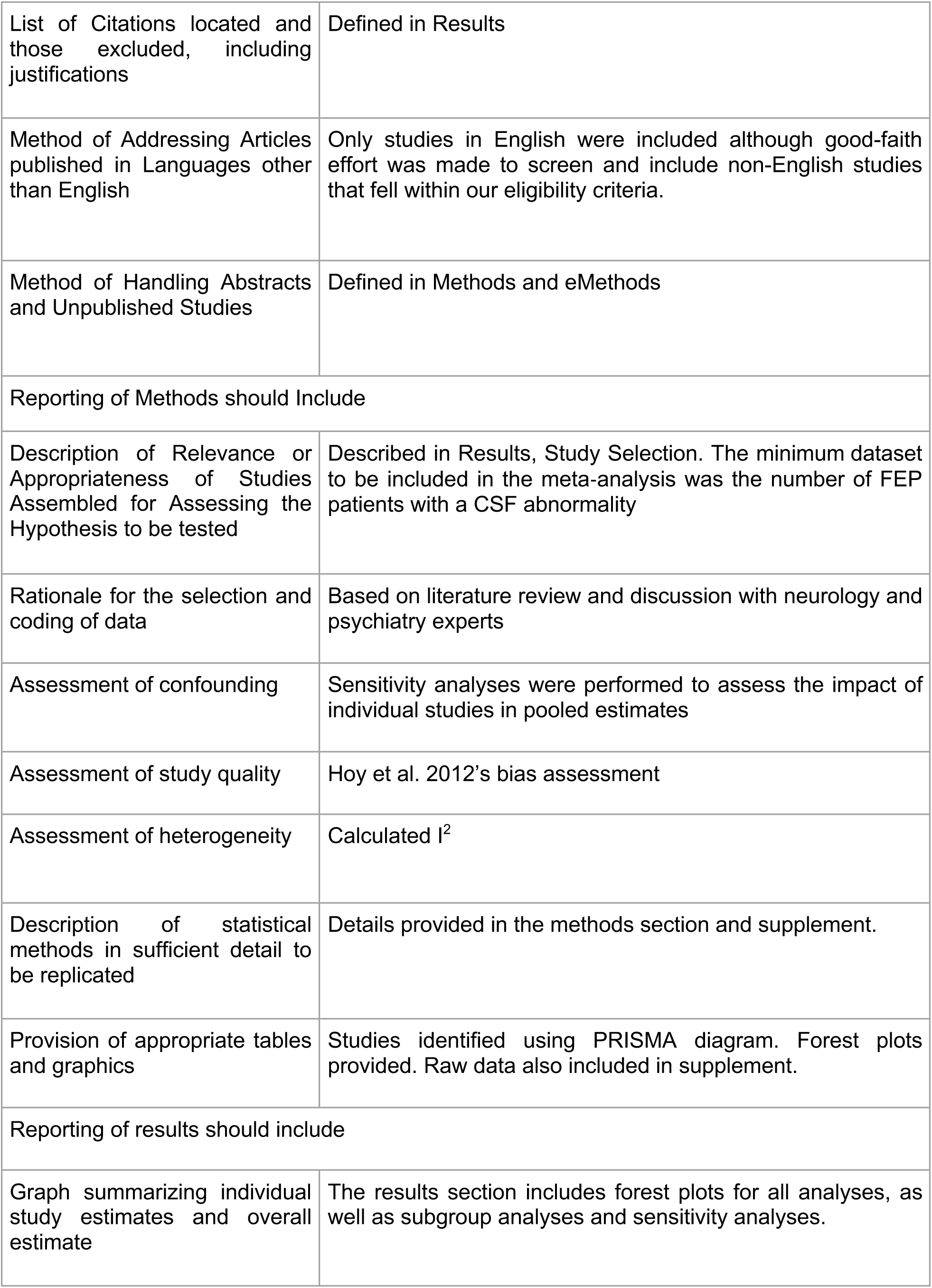
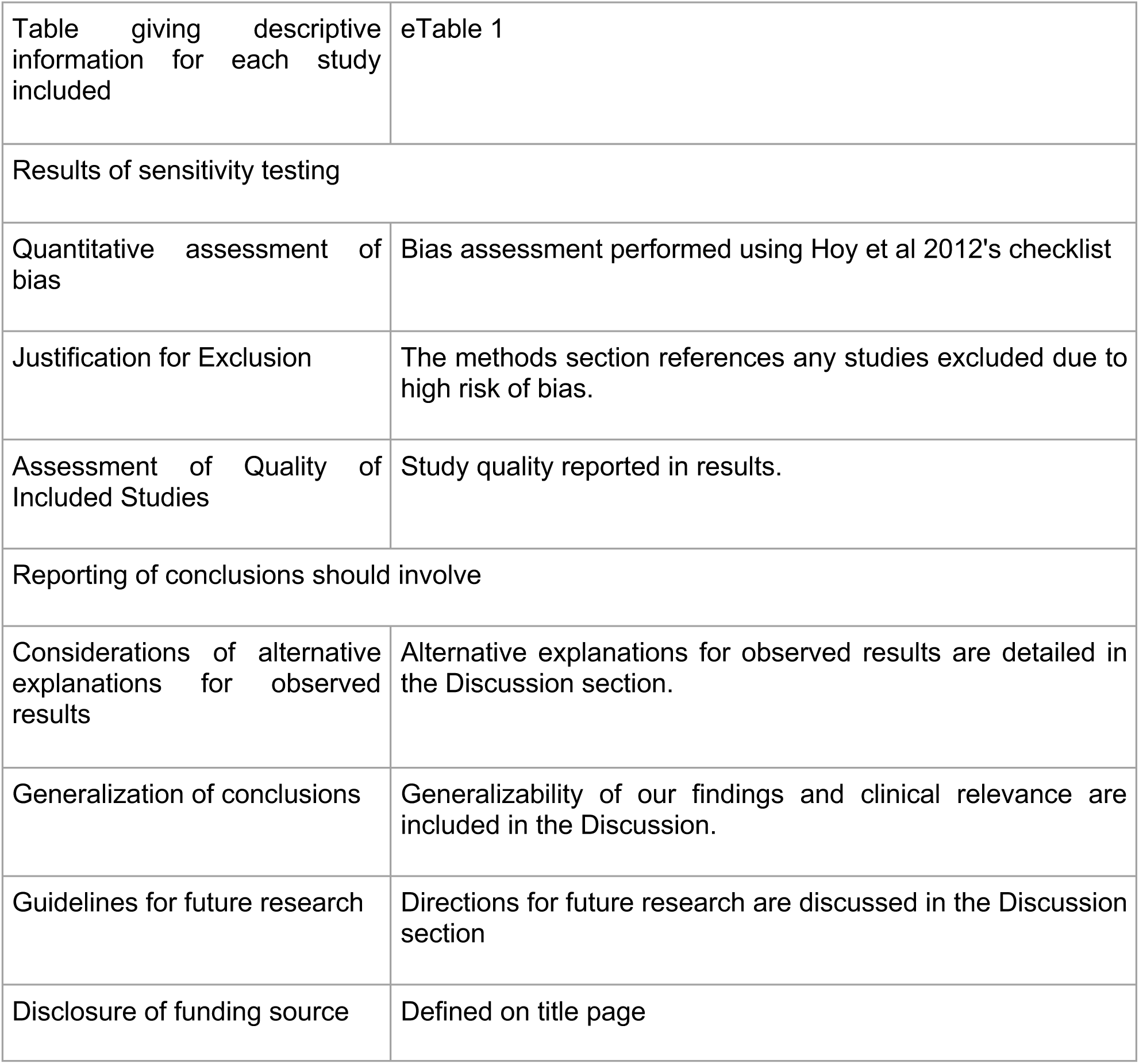
MOOSE Checklist

**eFigure 5.**
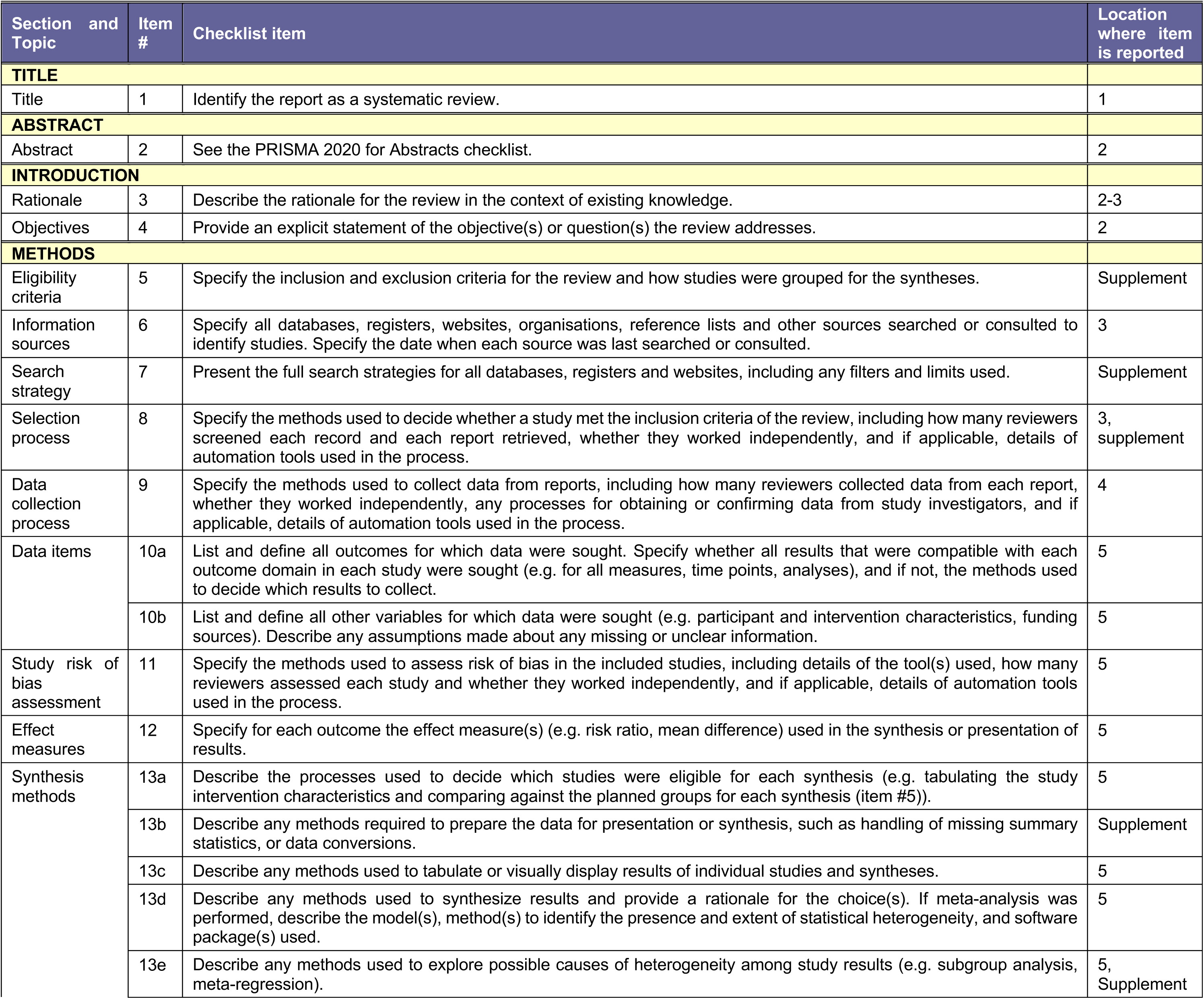
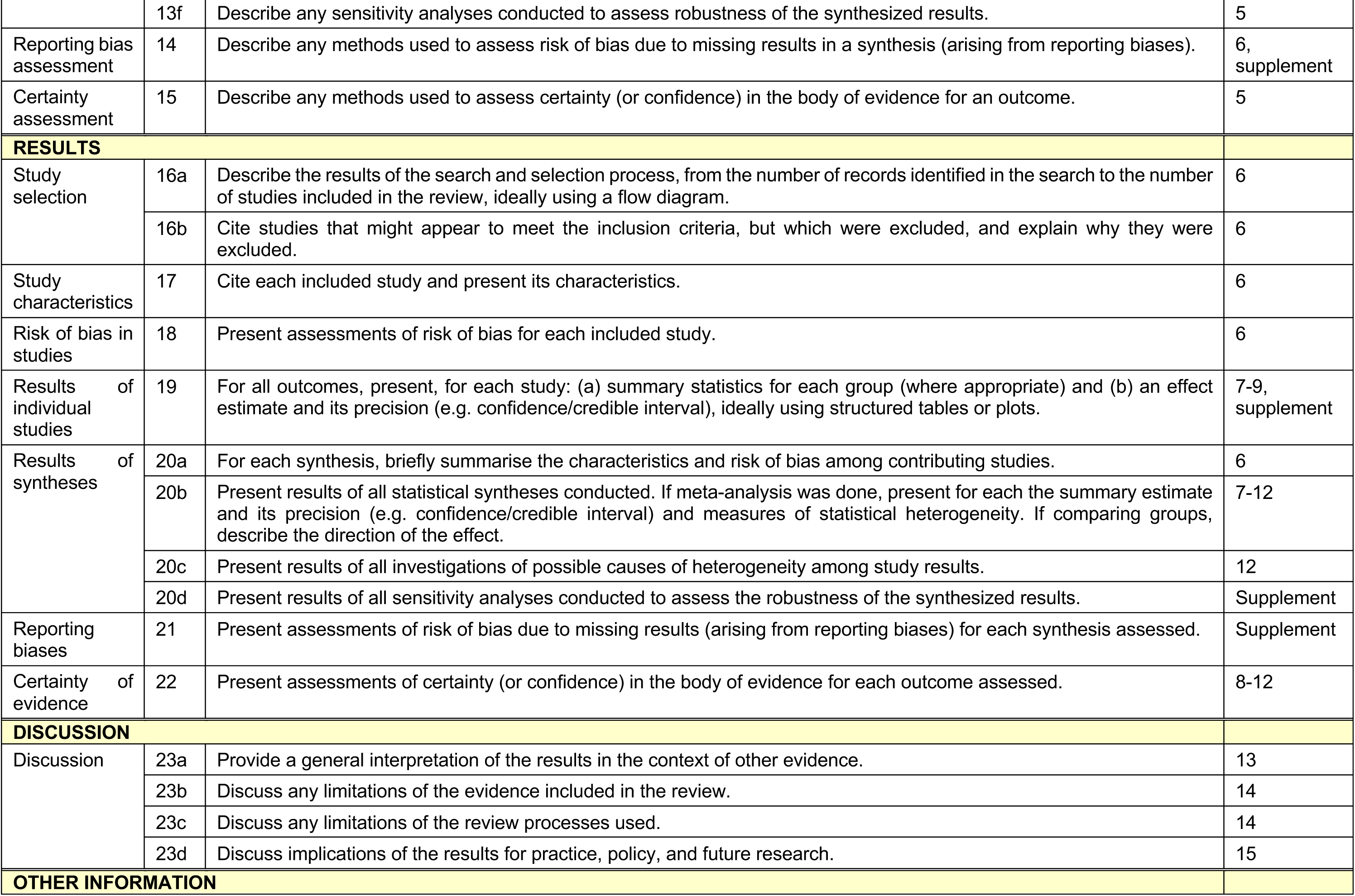
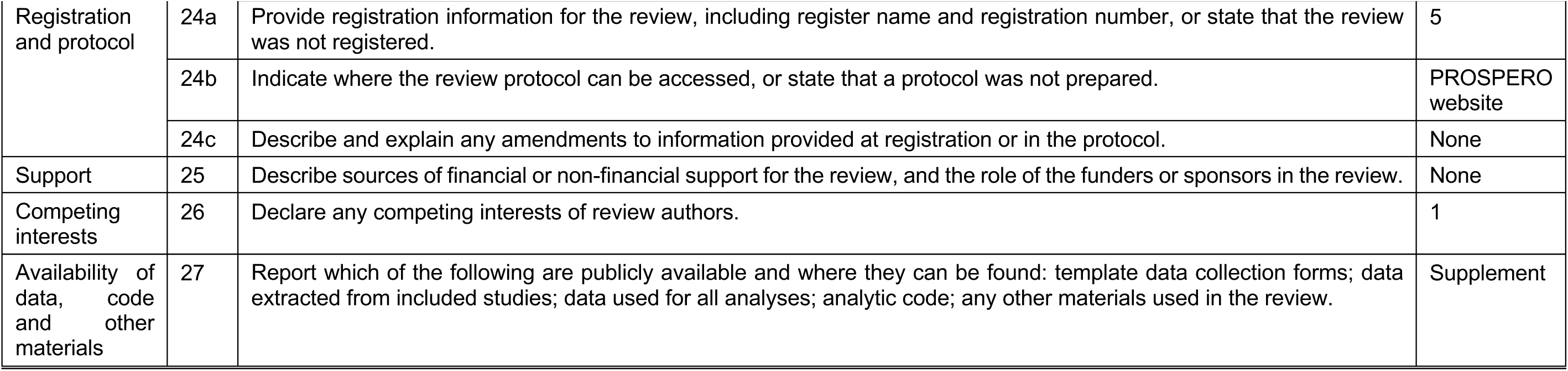
Prisma Checklist

**eTable 1.** Study inclusion/exclusion criteria*direct quotes are denoted by quotation marks.

| Study | Recruitment setting | Patient Inclusion Criteria |
| --- | --- | --- |
| Campana 2023 | patients at tertiary care hospital in Department of Psychiatry and Psychotherapy LMU University Hospital | "Medical records of patients suggesting clinical signs of a possible underlying autoimmune encephalitis (based on careful screening of all available data on clinical and physical examinations) were excluded from our analysis." |
| Campana 2022 | retrospective analysis from inpatients admitted to tertiary care hospital at Department of Psychiatry and Psychotherapy at LMU Munich selected for FEP patients | "Patients with clinical signs that could indicate the presence of an underlying autoimmune encephalitis (eg seizures, movement disorders, autonomic instability) were excluded based on thorough screening of all available data on clinical and physical examination." |
| Du 2023 | patients hospitalized in Departments of Neurology and Psychiatry of Sichuan Provincial People's Hospital | "Patients with mild or non-severe COVID-19 who tested positive for severe acute respiratory syndrome coronavirus 2 (SARS-CoV-2) (antigen or nucleic acid test) before or after admission; patients who presented abnormal mental behavior on admission, with delusions, hallucinations, and confusion in speech, behavior, and thinking as the main clinical manifestations." |
| Endres 2017 | patients at tertiary care hospital (Department of Psychiatry and Psychotherapy at University Medical Center Freiburg) | "Patients diagnosed with schizophreniform syndrome included those with schizophrenia (N=56), schizoaffective disorder (N=29), acute polymorphic psychotic disorder (N=14), and substance-induced psychosis (N=1). Only CSF/serum samples from patients with complete thyroid hormone status (thyroid-stimulating hormone [TSH], triiodothyronine [t3], and thyroxine [t4]) were included. For pragmatic reasons, we selected the first 100 patients since 2006. The study included 13 patients who underwent lumbar puncture between 2006 and 2009 (i.e., CSF analysis was performed if there were signs of possible neuroinflammatory features). The other 87 patients underwent lumbar puncture since June 2009, when CSF analysis was offered as a routine procedure." |
| Endres 2020 | patients at tertiary care hospital (Department of Psychiatry and Psychotherapy at University Medical Center Freiburg) | "All inpatients with schizophreniform syndromes (according to the International Statistical Classification of Diseases and Related Health Problems criteria, unipolar depression following ICD-10: F32.X, F33.X, F06.3 and bipolar disorder following ICD- 10: F30.X, F31X, F06.3) who underwent an LP at the Department of Psychiatry and Psychotherapy were included. Other preexisting (e.g., earlier stroke) or newly described (e.g., migraine) neurological comorbidities were recorded but not considered as an exclusion criteria if the LP was performed within the diagnostic process of the psychiatric disorder." |
| Endres 2015 | university clinic of psychiatry and psychotherapy Freiburg | "From 2006 to 2013, we acquired 180 CSF samples from psychotic patients. Between 2006 and 2009, CSF examinations were only performed in cases in which organic brain disease was suspected." |
| Endres 2024 | all adult patients treated at specialized ward for psychosis and secondary mental disorders at University of Freiburg | "All adult patients treated in our specialized ward for psychosis and secondary mental disorders for over 2.5 years were screened, and those with primary or secondary psychotic syndromes were included in the current analysis (schizophreniform syndromes: F20.X, F29.X, F06.02, F10.5, F19.5; depression with psychotic symptoms: F32.3, F33.3; mania with psychotic symptoms: F30.2; bipolar affective disorder, currently manic or major depressive episode with psychotic symptoms: F31.2, F31.5). Thus, patients with secondary psychoses or rheumatological diseases were not excluded." |
| Gine-Serven 2022 | acute inpatient units (adult or child and adolescent units) from the Department of Mental Health at the Hospital of Mataró between 1 June 2018 and 31 March 2020 were offered to participate in the study | Patients with FEP who were admitted to acute inpatient units at the Hospital of Mataró between 1 June 2018 and 31 March 2020. |
| Guasp 2021 | Hospital Clínic Institut d'Investigacions Biomèdiques August Pi i Sunyer (IDIBAPS), University of Barcelona | "All patients with FEP admitted to 2 hospitals at University of Barcelona, between June 1, 2018, and March 31, 2020, were invited to participate in the study. FEP was defined as new-onset disorganized behavior accompanied by delusions or hallucinations." |
| Hinotsu 2022 | newly hospitalized patients at Okayama Psychiatric Hospital initially diagnosed with schizophrenia, mood disorder, epilepsy, psychiatric symptoms | "Patients diagnosed with schizophrenia, a mood disorder or epilepsy with psychiatric symptoms" |
| Kelleher 2020 | pts recruited from general psychiatric services from catchment areas strictly defined as attached to St. James's hospital and Tallaght University Hospital in Dublin and Newcastle hospital Wicklow. FEP cases were recruited via community mental health teams who were educated about study and asked to recruit patients who met inclusion criteria. liaised with neurology centres to identify any additional cases diagnosed with NMDA-R Ab encephalitis within catchment area. Treatment resistant patients were invited by nursing staff from Clozapine clinics within catchment area | "Inclusion criteria for the FEP cohort comprised of being resident in the study catchment area for longer than six months based on their address within the catchment area, being aged between 18 and 65 years old and to be experiencing their first episode of psychosis. Only 4 seropositive cases had an LP." Excluded: Presence of known pre-existing neurological disorder. |
| Kruse 2015<br>ANTI-NMDAR IgG seropositive | NMDA receptor IgG seropositive patients seen at Mayo Clinic rochester from 2009-2013 | Group 2: NMDA receptor IgG. Retrospective assessment for each seropositive patient. "High likelihood assigned when new psychiatric symptoms of subacute onset were accompanied by at least one of the following: ew-onset movement disorder, encephalitic-appearing magnetic resonance imaging (MRI), encephalopathic or epileptiform electroencephalogram (EEG), inflammatory cerebrospinal fluid (CSF) (with protein exceeding 70 mg/dL, pleocytosis, or at least 4 CSF-exclusive oligoclonal bands), neural autoantibody at least 50% predictive for cancer (eMethods), or past episode of immunotherapy-responsive encephalitis." |
| Kruse 2015 | adult psychiatric inpatients at Mayo clinic who had comprehensive serological evaluation for neural autoantibodies | Group 1: adult psychiatric inpatients whose initial neuropsychiatric evaluation included comprehensive serologic evaluation for neural autoantibodies (January 1, 2002 - December 31, 2011) |
| Maurus 2023 | FEP patients admitted to tertiary care hospital to Department of Psychiatry and Psychotherapy at University Hosital, Ludwig Maximilian University of Munich Germany | FEP admitted between 1 Jan 2008 and 1 Aug 2018 -Inclusion criteria were that patients have a main diagnosis of F2x (schizophrenia spectrum according to the ICD-10) in the discharge summary of the first in-patient stay, and that patients had undergone both a lumbar puncture and neurocognitive testing within the clinical routine assessments before receiving this diagnosis. In our hospital, CSF analyses are usually offered to every FEP patient by the treating physicians so as to rule out organic psychoses, according to best clinical practice. "We also included patients who initially presented themselves with psychotic symptoms and a non-F2x-diagnosis (e.g. drug-induced psychosis), and were later diagnosed within the schizophrenia spectrum. any psychiatric or physical pre- diagnoses possibly resulting in cognitive impairments or BBB dysfunction (such as intellectual disability, perinatal asphyxia or stroke); abnormal brain magnetic resonance imaging findings suggestive of a relevant co-occurring CNS disorder (e.g. neoplasms); and a time interval of more than 21 days between CSF and blood sampling, or of more than 90 days between CSF sampling and neurocognitive testing, to ensure a clinically meaningful temporal association. Based on thorough screening of all available data, 45 patients were excluded before data analysis." |
| Oviedo-Salcedo 2018 | patients with schizophrenia who underwent a lumbar puncture through Department of Psychiatry and Psychotherapy at the Klinikum der Universitat Munchen | "We included patients with documented ICD-10 diagnoses of schizophrenia spectrum disorders and excluded patients with a diagnosis of organic psychotic disorders (F06.0/F06.1/F06.2), drug-induced psychotic disorder (F1X.5) or clinical signs that could indicate the presence of an underlying autoimmune encephalitis(e.g. seizures, movement disorders, autonomic instability), based on thorough screen- |
|  |  | ing of all available data on clinical and physical examination. excluded patients with a diagnosis of organic psychotic disorders (F06.0/F06.1/F06.2), drug-induced psychotic disorder (F1X.5) or clinical signs that could indicate the presence of an underlying autoimmune encephalitis<sup>7</sup> (e.g. seizures, movement disorders, autonomic instability), based on thorough screening of all available data on clinical and physical examination.” |
| Restrepo-Martinez 2020 | Patients were recruited via in- and outpatient clinics in the Capital Region of Denmark, whereas healthy controls were recruited via a Danish website (ForsÃfÂgsperson.dk).<br>Patients and controls came from the same community. All participants had to be aged 18-50 years, and patients were to have been diagnosed with a nonorganic, non-affective psychotic disorder (International Classification of Diseases, Tenth Revision (ICD10) F20/F22-29) within a year prior to inclusion. | “Patients who had not received a CSF analysis in the past as part of the diagnostic work-up. Patients with organic psychotic disorders (ICD-10: F06.0/F06.1/F06.2) and substance-induced disorders (ICD-10: F1X.5) were excluded from our analyses in order to isolate cases of primary psychosis as defined by clinical judgment. We also screened for major neurological symptoms in the clinical documentation such as epileptic seizures and disturbed consciousness that could indicate a major neurological disease at the time of admission. The decision to perform the lumbar puncture was made by the treating physicians according to best clinical practice. In our psychiatric hospital, CSF analyses are usually offered to every FEP patient and to MEP patients who had not received a CSF analysis in the past as part of the diagnostic work-up.” |
| Runge 2022 | patients with schizophrenia spectrum disorders who were treated as inpatients in the Department of psychiatry and psychotherapy of the medical center of the University of Freiburg | Exclusion criteria: known substance use disorders and immunological disorders known for brain involvement. |
| Jeppesen 2022 | Patients were recruited via in- and outpatient clinics in the Capital Region of Denmark, whereas healthy controls were recruited via a Danish website (ForsÃfÂgsperson.dk). | Patients and controls came from the same community. All participants had to be aged 18-50 years, and patients were to have been diagnosed with a non organic, non-affective psychotic disorder (International Classification of Diseases, Tenth Revision (ICD10) F20/F22-29) within a year prior to inclusion. |
| Runge 2021 | patients with schizophrenia spectrum disorders who underwent LP during acute inpatient stay at University of Freiburg | Patients with Schizophrenia Spectrum Disorders who underwent lumbar puncture during their acute inpatient stay to exclude secondary organic causes of the disorder. They had to have received prior MRI imaging and be able to give consent. Patients with schizophrenia (ICD-10: F20), delusional disorders (ICD-10: F22), brief psychotic disorder (ICD-10: F23) and schizoaffective disorders (ICD-10: F25) were included. Patients with neurodegenerative diseases, acute clinical infections, or current substance dependence disorders, as well as neurological or immunological diseases with known brain involvement were excluded from the study. |
| Kranaster 2011 | first episode schizophrenia or schizophreniform patients at University of Cologne according to DSM-IV criteria | no known previous or current treatment with antipsychotic medication |
| Steiner 2013 | acutely ill patients from scientific blood bank at Magdeburg University Department of Psychiatry | “Patients with psychosis resulting from other medical conditions and substance-induced psychosis were excluded by a thorough physical examination, routine blood analysis (including differential blood cell count, kidney function tests, and levels of C-reactive protein, glucose, lipids, liver, enzymes, and thyroid hormones), screening for illegal drugs,<br>And magnetic resonance imaging the brain. In Addition,electroencephalography (EEG)was performed.The same tests were carried out in the control, MD, and BLPD participants.” |
| Srikanth 1994 | patients admitted to NiMHANS Bangalore | “They met the exclusion criteria: absence of poor nutrition (based on general physical examination), anemia, generalized bacterial or fungal infections, recent trauma, alcohol and drug abuse, neurological disorders, therapeutic drugs (including psychotropic drugs) during the last 6 months. They never had organ transplantation, elec-electroconvulsive therapy (ECT), or recent vaccination or blood transfusion.” |
| Seppala 2014 | geriatric psychiatry department of Helsinki University Central Hospital consisting of 4 wards | “Psychotic symptoms had not lasted more than 2 years before their admission.No patients with a known diagnosis of dementia (AD, DLB, etc.) at admission were included. Patients with other diseases of the central nervous system (e.g., Parkinson disease [PD] or major stroke) were also excluded.” |
| Bien 2021 | pts recruited from two clinical sites with community service obligations for inpatient and outpatient services, including early recognition for psychosis units in Germany, Department of Psychiatry and Psychotherapy at University of Cologne, Department of Psychiatry and Psychotherapy at Central Institute of Mental Health Mannheim | “Individuals met the following criteria: attenuated positive symptoms, brief, limited intermittent psychotic symptoms that spontaneously resolved within a week, a recent decline in function.” |
| Bauer 1987 | patients were investigated at Department of Neurology and Psychiatry at Bundeswehrkrankenhaus Ulm | Only included patients newly diagnosed as schizophrenic |
| Rauber 2021 | retrospective screening of local clinical patient database at University Clinic Munster, Germany to identify patients with available CSF data and primary psychotic disorders encompassing schizophrenia | Identified patients with available CSF mFC data and primary psychotic disorders encompassing schizophrenia, delusional disorders, acute and transient psychotic disorders or schizoaffective disorders. |
| Tang 2019 | first episode psychosis patients recruited from institute of mental health, national neuroscience institute, a neurological center, and Tan Tock Seng Hospital, a general hospital in Singapore between September 2011 and October 2013 | Inclusion criteria were age 16–50 years, the presence of less than 1 month of psychotic symptoms, and any of the following neurological symptoms: cognitive dysfunction, seizures, abnormal movements, or autonomic dysfunction. Patients were excluded if there was alcohol or drug-induced psychosis as determined by clinical history and/or a positive drug screen. |
| Rattay 2021 | inpatients with psychosis with CSF analysis between 01/2016 and 06/2020 at Department of Psychiatry and Psychotherapy at University Hospital Tubingen Germany | Patients on any four of the acute admission wards or two wards specialized in the psychotherapy of psychotic disorders |
| Maier 2024 | inpatients treated at the Department of Psychiatry, Social Psychiatry and Psychotherapy at Hannover Medical School | Patients with a previously diagnosed neurological or organic mental disorder (ICD-10: F00-F09), the second LP of the same patient, and CSF analysis that was not indicated for the differential diagnostic work-up of psychiatric symptoms were excluded. |
| Muller 1995 | schizophrenic patients from 18 to 57 at the Psychiatric Hospital at University of Munich | Patients presenting a clinical indication for an LP, e.g. the first detailed investigation in order to exclude an organic disease, the first hospitalization, the first onset; first detailed investigation in order to exclude an organic disease, the first hospitalization |
| Theorell 2021 | first episode patients were recruited from psychiatric wards and outpatient departments as a part of the Karolinska Schizophrenia Project | First-episode psychosis patients (n = 74), defined as first physician contact due to psychotic symptoms. Exclusion: overt abnormal signs on neurological examination, seizures, autonomic instability or a movement disorder. |
| Soderland |  | first-episode |
| Torrey 1978 | first admission schizophrenia patients, multi-episode schizophrenia, schizo-affective psychosis, and manic depressive psychosis amongst inpatients at National Institute of Mental Health Bethesda. Controls were patients at National Naval medical center | “The patient group was those with first admission schizophrenia, multiple admission schizophrenia, schizo-affective psychosis and manic depressive psychosis. One control group were surgical patients with no known neurological or psychiatric disease. Finally there was another comparison group of 80 patients with neurological disease.” |

**eTable2.** Study Characteristics.

| Study | Year | Country | FEP (n) | Control (n) | Age (years) | Female (percent) | Psychosis Duration (weeks) | Quality Assessment |
| --- | --- | --- | --- | --- | --- | --- | --- | --- |
| Campana et al. | 2023 | Germany | 222 | NA | 34.89 | 46.4 |  | 8 |
| Campana et al. | 2022 | Germany | 314 | NA | 35.1 | 45.2 | 52.4 | 6 |
| Du et al. | 2023 | China | 57 | NA |  |  |  | 6 |
| Endres et al. | 2017 | Germany | 100 | NA | 33.32 | 59 |  | 6 |
| Endres et al. | 2020 | Germany | 279 | NA | 32.72 | 54 |  | 8 |
| Endres et al. | 2015 | Germany | 33 | NA |  |  |  | 7 |
| Endres et al. | 2024 | Germany | 161 | NA | 34.71 | 56 |  | 6 |
| Gine-Serven et al. | 2022 | Spain | 95 | NA | 35 | 39 |  | 9 |
| Guasp et al. | 2021 | Spain | 105 | NA | 30 | 42 |  | 8 |
| Hinotsu et al. | 2022 | Japan | 151 | NA |  |  |  | 9 |
| Kelleher et al. | 2020 | Ireland | 112 | NA | 33 | 38 |  | 7 |
| Kruse et al. | 2015 | USA | 13 | 173 | 27 | 92 |  | 6 |
| Maurus et al. | 2023 | Germany | 121 |  | 35.1 | 50 | 48.8 | 7 |
| Oviedo-Salcedo et al. | 2018 | Germany | 332 |  | 36.85 | 46.3 | 246.32 | 3 |
| Oviedo-Salcedo et al. | 2021 | Germany | 128 |  | 34.43 | 48.4 | 44.68 | 8 |
| Restrepo-Martinez et al. | 2020 | Mexico | 24 |  | 24.5 | 38.3 | 46.4 | 8 |
| Runge et al. | 2022 | Germany | 100 | Pts with IIH | 33.7 | 60 |  | 8 |
| Jeppesen et al. | 2024 | Germany | 104 |  | 26.1 | 38.5 | 86.8 | 7 |
| Runge et al. | 2021 | Germany | 40 |  | 34.6 | 60 |  | 7 |
| Kranaster et al. | 2011 | Denmark | 155 |  | NA | 0 |  | 8 |
| Steiner et al. | 2013 | Germany | 229 | 30 healthy controls | NA | 0 |  | 6 |
| Srikanth et al. | 1994 | India | 35 | 35 controls (women undergoing epidural) | NA | 0 |  | 6 |
| Seppala et al. | 2014 | Finland | 51 |  | NA | 78 |  | 6 |
| Bien et al. | 2021 | Germany | 103 | Yes, healthy controls 40 | NA | 19.1 |  | 9 |
| Bauer et al. | 1987 | Germany | 15 |  | NA | 0 |  | 6 |
| Rauber et al. | 2021 | Germany | 59 | Control group with IIH 52 pts | NA | 37 |  | 8 |
| Tang et al. | 2019 | Singapore | 15 |  | NA | 80 |  | 7 |
| Rattay et al. | 2021 | Germany | 144 |  | NA |  |  | 9 |
| Maier et al. | 2024 | Germany | 254 |  | 33.8 | 56.9 | 103.84 | 8 |
| Muller et al. | 1995 | Germany |  |  |  |  |  | 7 |
| Theorell et al. | 2021 | Sweden and UK |  |  |  |  |  | 8 |
| Soderland et al. | 2009 | Sweden |  |  |  |  |  | 8 |
| Torrey et al. | 1978 | USA |  |  |  |  |  | 6 |
| Lopes et al. | 2024 | Portugal | 68 |  | 49 | 60.3 |  | 7 |
| Coughlin et al. | 2016 | USA | 14 |  | 24 | 21 | 803 | 8 |

**eTable 3.**
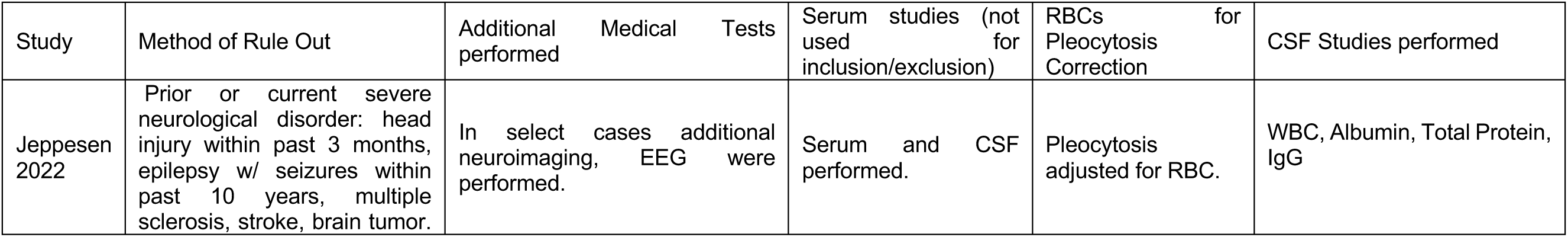
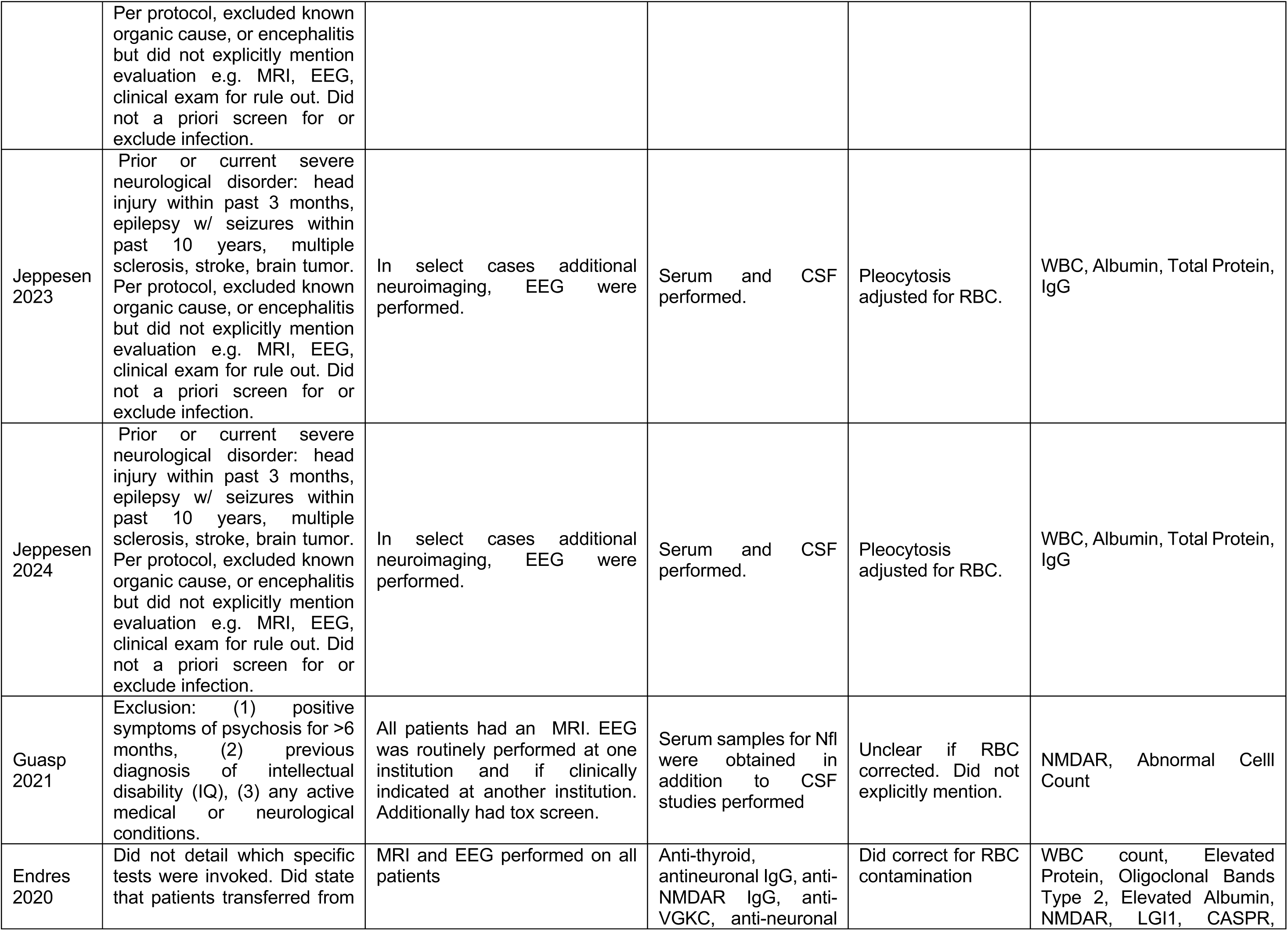
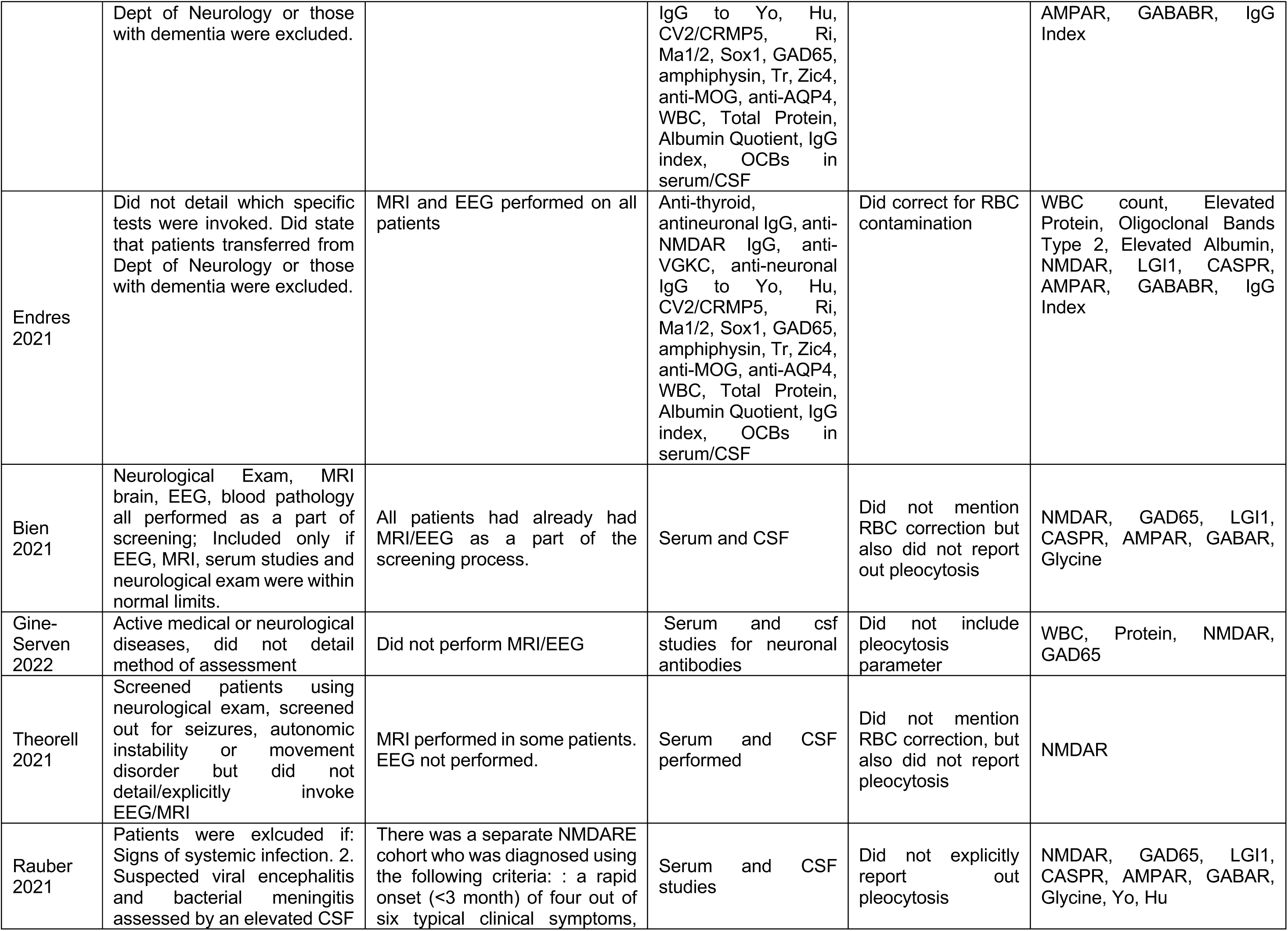
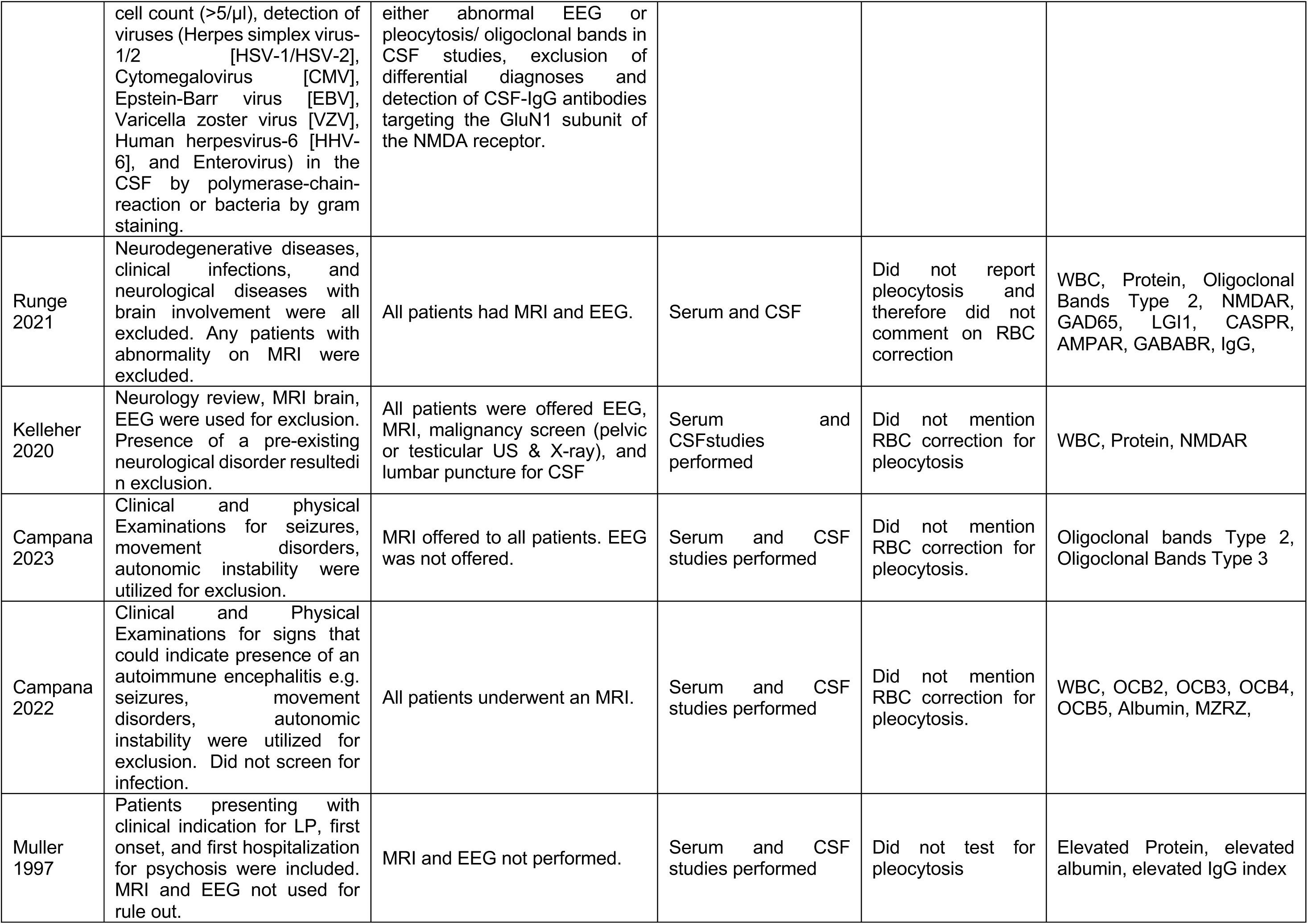
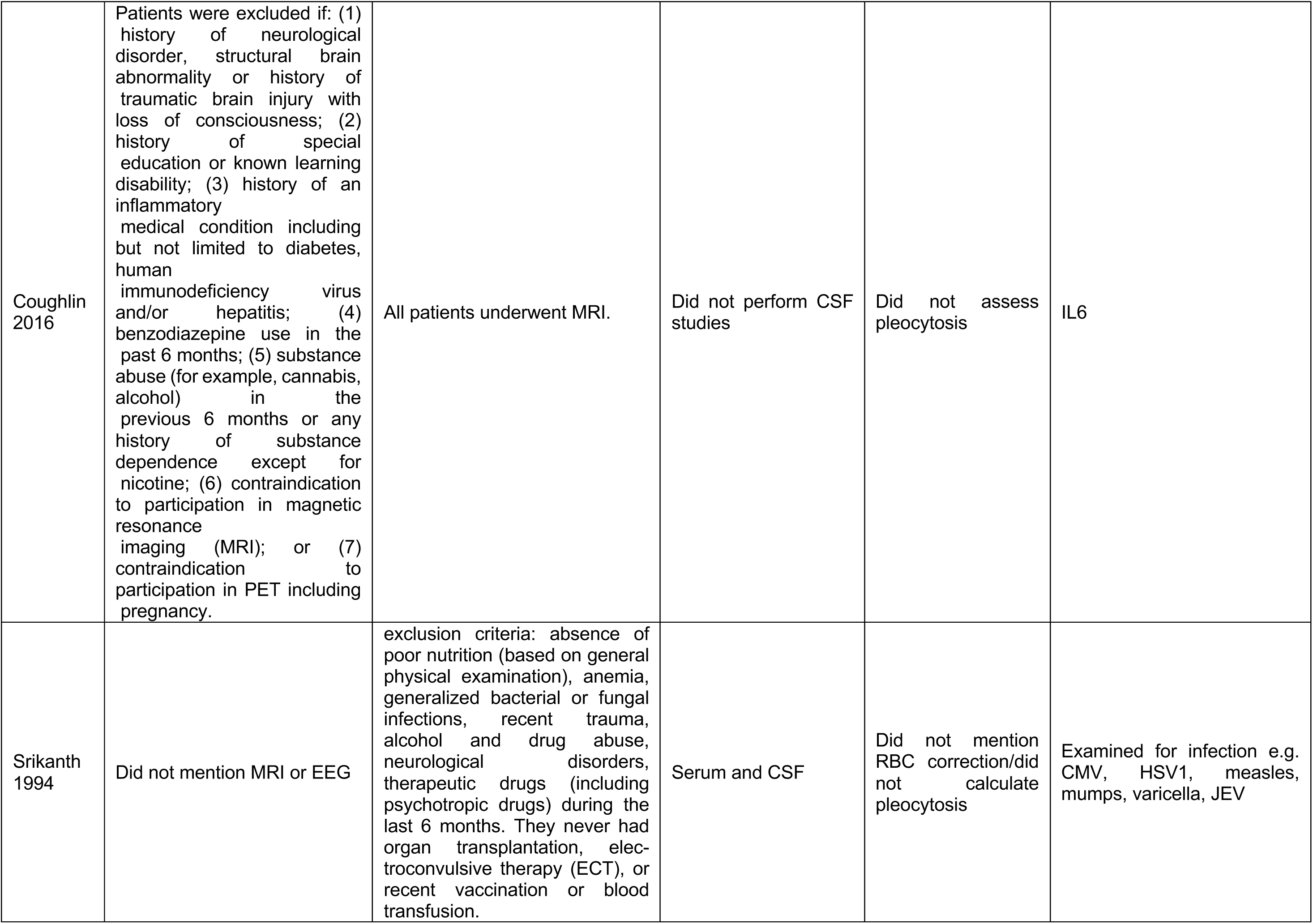
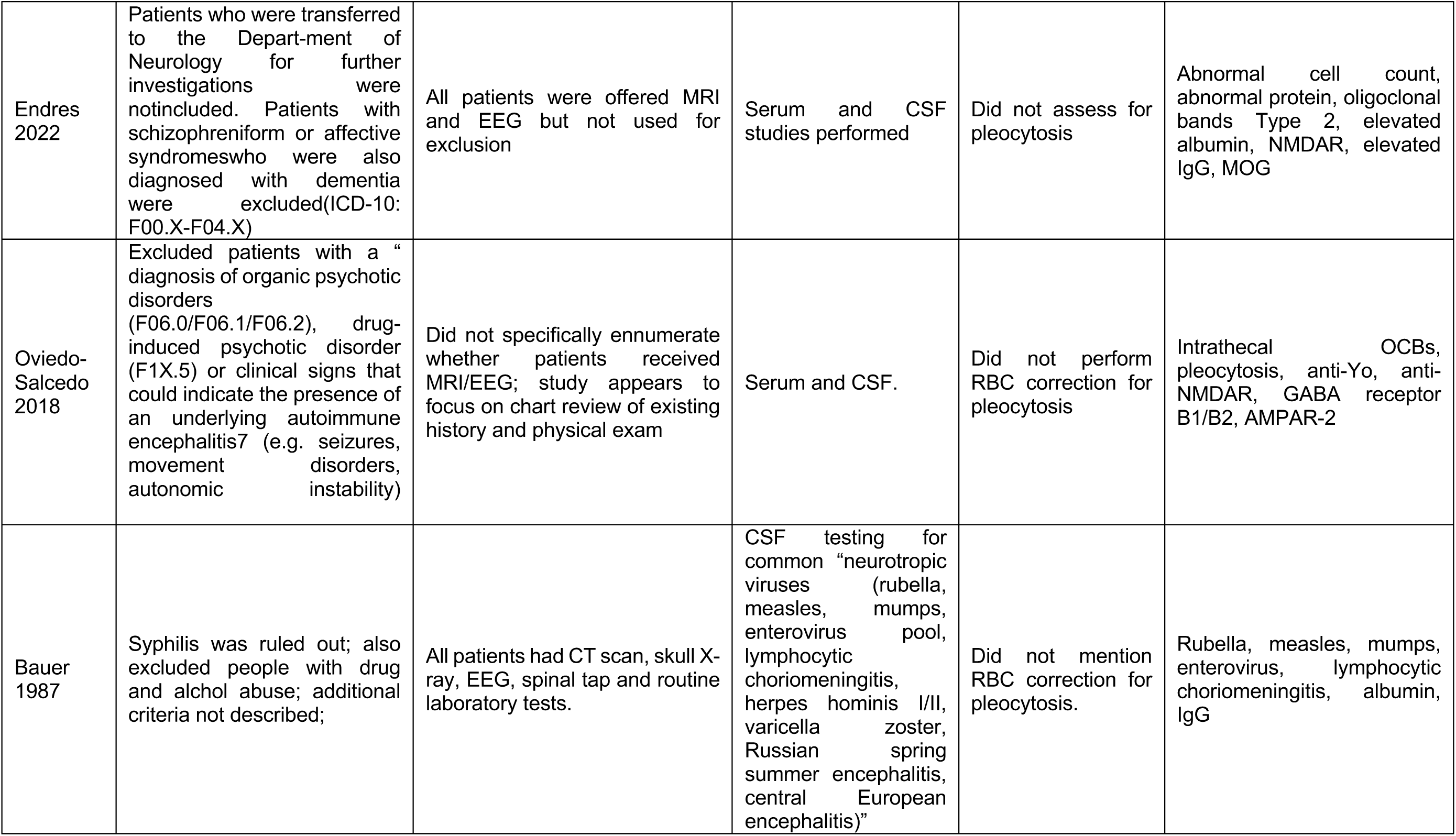
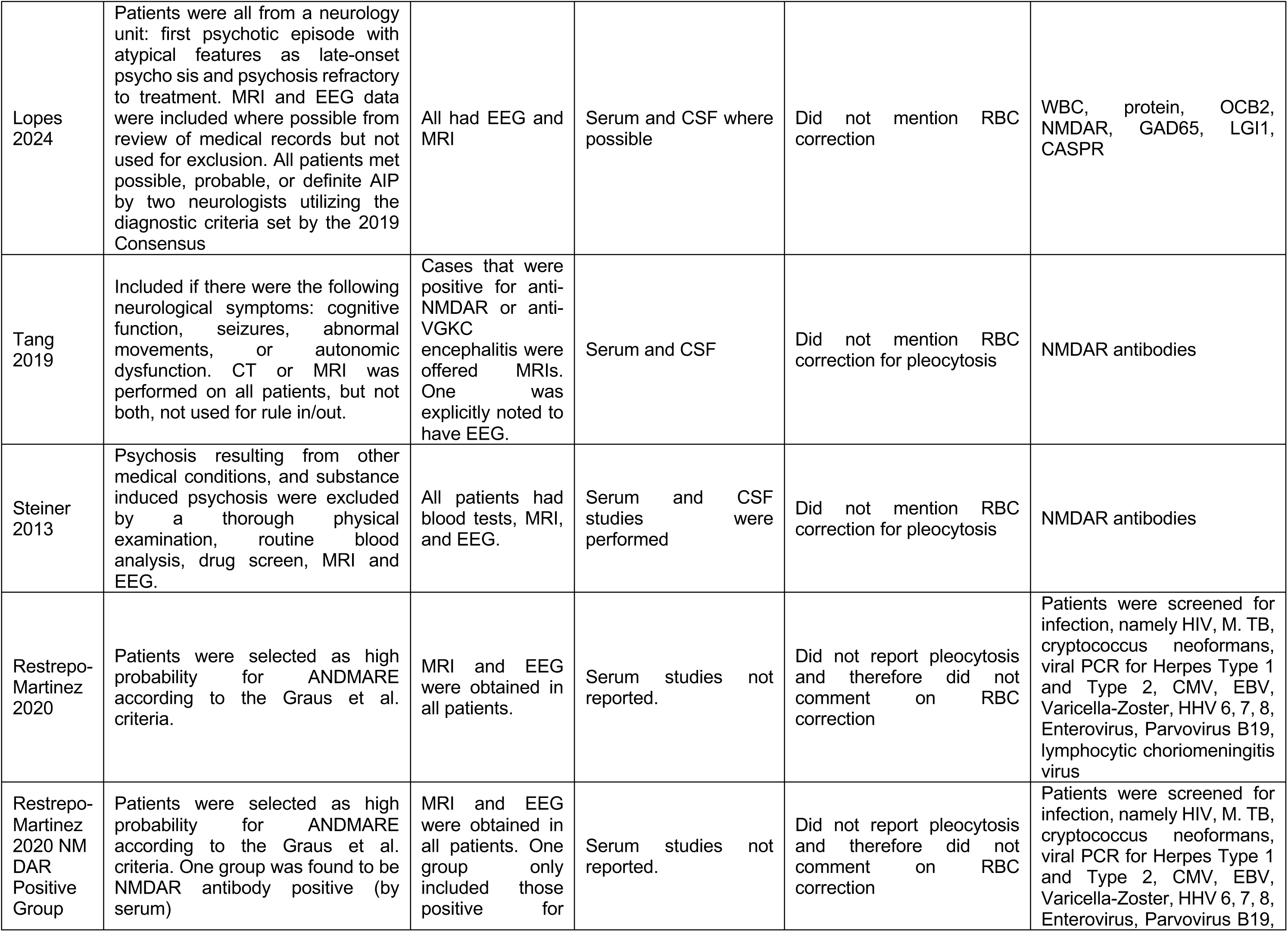
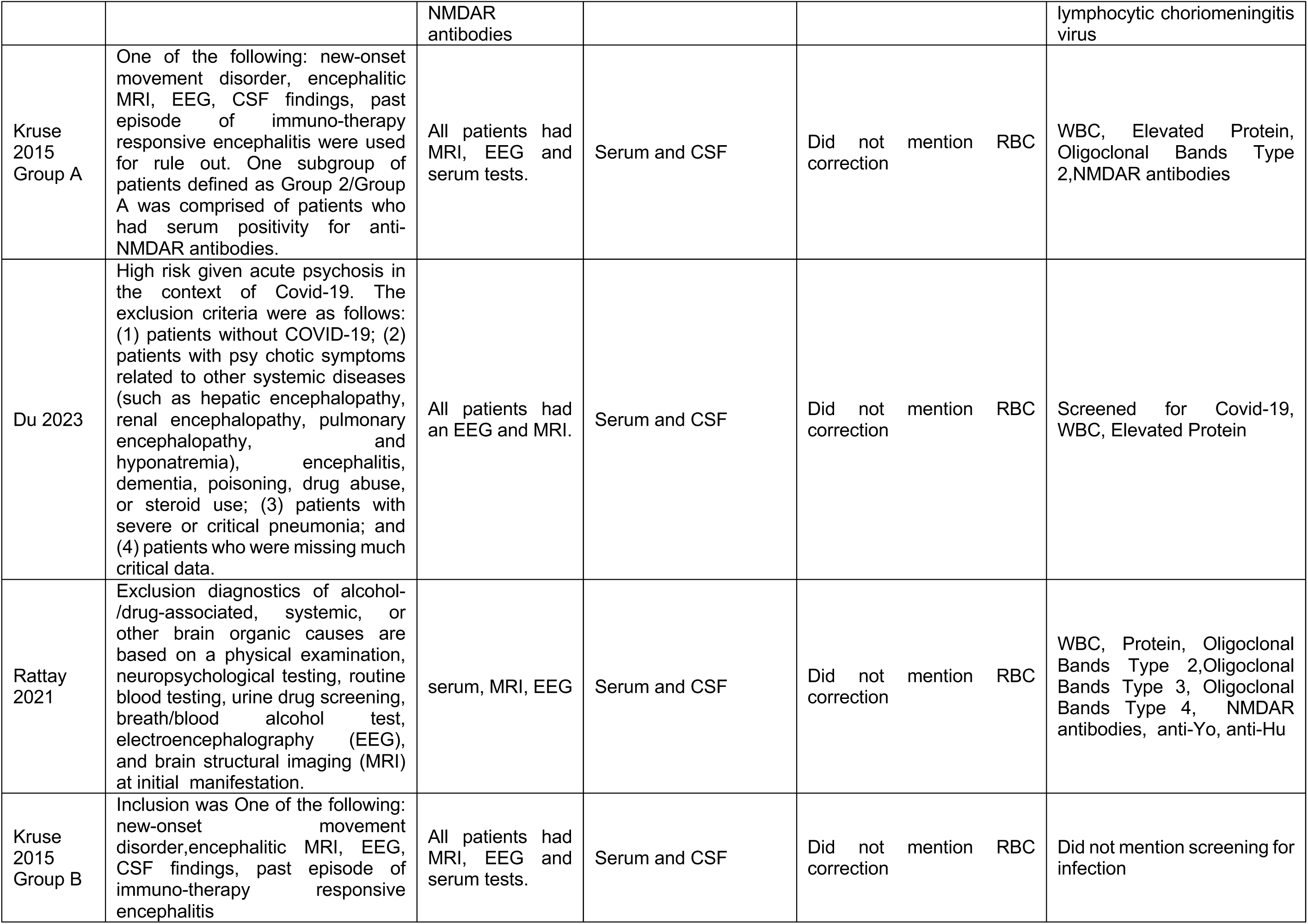
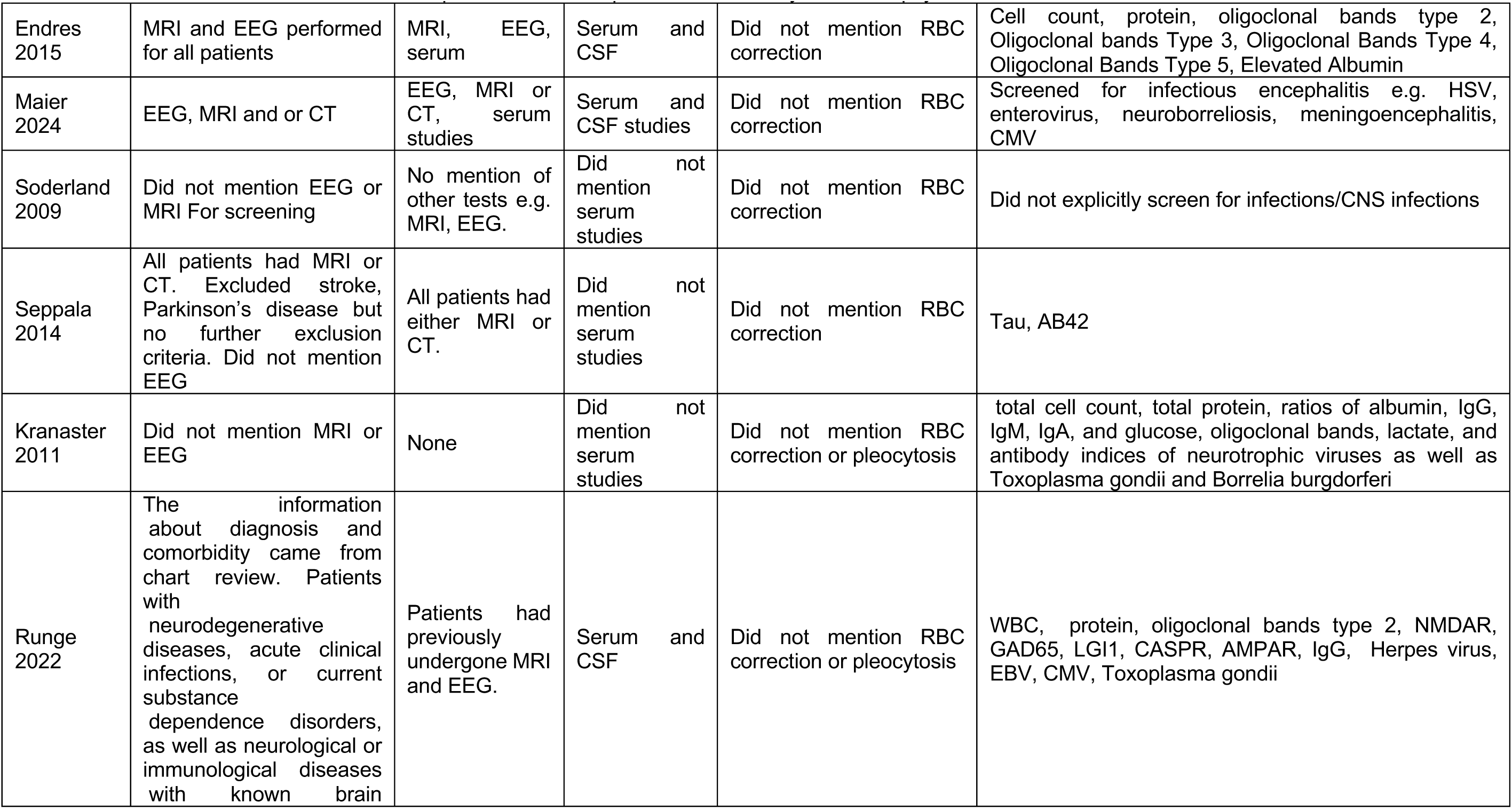
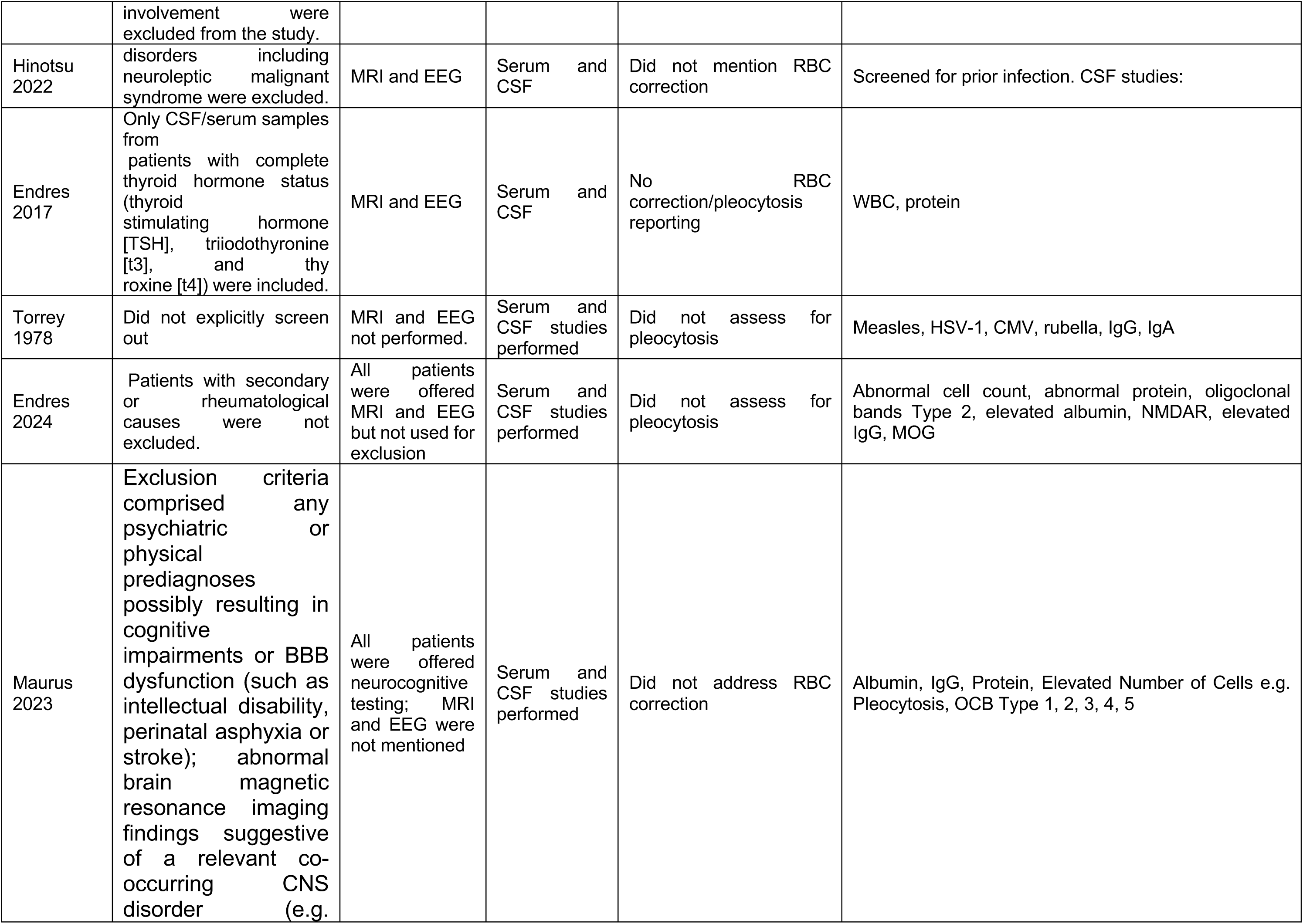
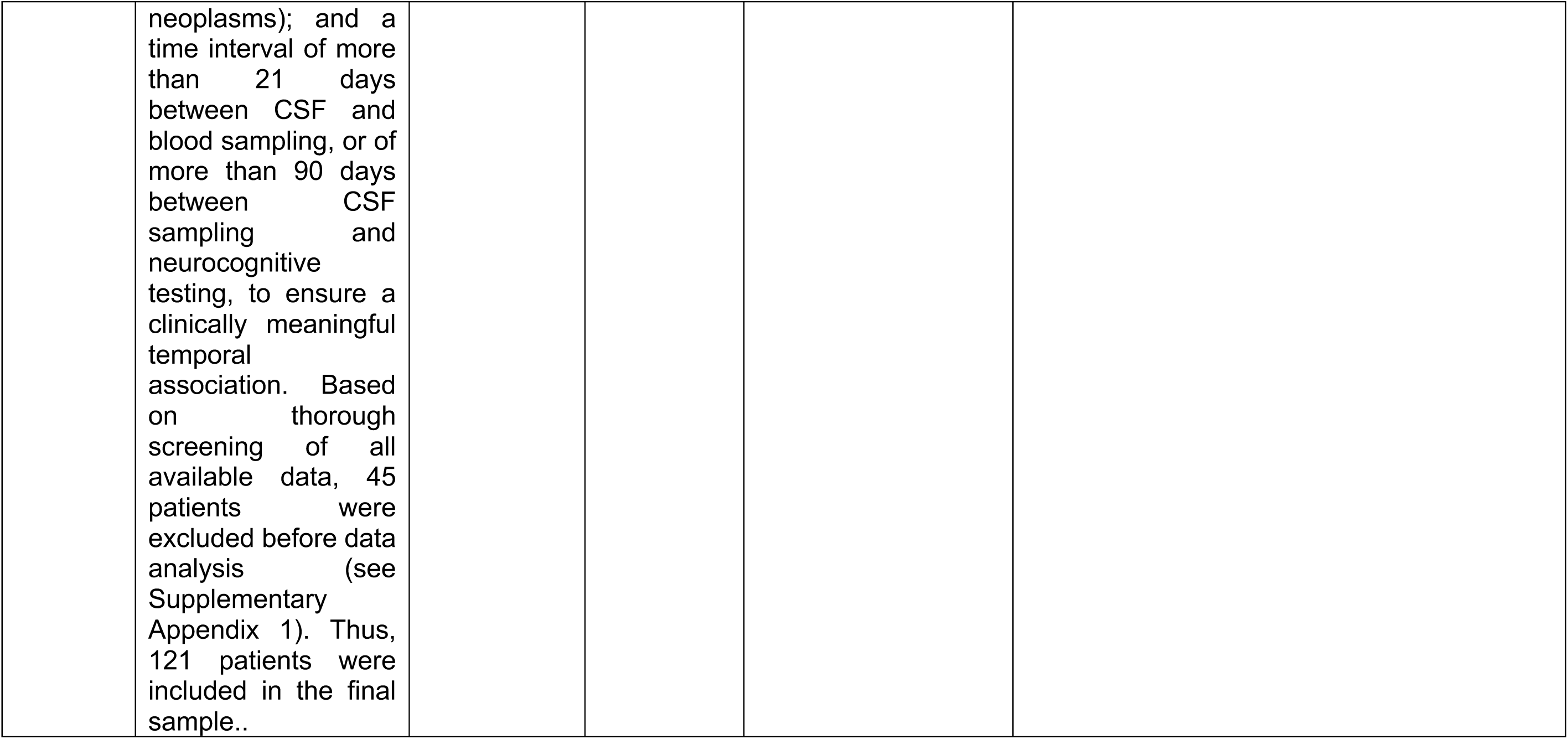
Protocol for how studies identified subgroups based off clinical suspicion for secondary causes of psychosis.

## Notes

### Competing Interest Statement

MLB receives royalties from Oxford University Press for his book, The Neuroethics of Biomarkers: What the Development of Bioprediction Means for Moral Responsibility, Justice, and the Nature of Mental Disorder. LD, GB, GM, MEB have no disclosures

### Funding Statement

The study did not receive any dedicated funding.

### Author Declarations

Electronic databases Ovid, Medline, Embase, PsychoINFO, and Web of Science were searched from inception to October 2024 for studies that met our inclusion criteria. References of included articles were also screened. See methods for full details.

### Summary of Updates

Prevalence estimates after identifying that an additional set of papers were derived from the same cohort.

